# Interplay of Race and Neighborhood Deprivation on Ambulatory Blood Pressure in Young Adults

**DOI:** 10.1101/2023.09.11.23295160

**Authors:** Soolim Jeong, Braxton A. Linder, Alex M. Barnett, McKenna A. Tharpe, Zach J. Hutchison, Meral N. Culver, Sofia O. Sanchez, Olivia I. Nichols, Gregory J. Grosicki, Kanokwan Bunsawat, Victoria L. Nasci, Eman Y. Gohar, Thomas E. Fuller-Rowell, Austin T. Robinson

## Abstract

**Background:** Ambulatory blood pressure (**BP**) monitoring measures nighttime BP and BP dipping, which are superior to in-clinic BP for predicting cardiovascular disease (**CVD**), the leading cause of death in America. Compared with other racial/ethnic groups, Black Americans exhibit elevated nighttime BP and attenuated BP dipping, including in young adulthood. Social determinants of health contribute to disparities in CVD risk, but the contribution of neighborhood deprivation on nighttime BP is unclear. Therefore, we examined associations between neighborhood deprivation with nighttime BP and BP dipping in young Black and White adults.

**Methods:** We recruited 21 Black and 26 White participants (20 M/27 F, mean age: 21 years, body mass index: 25±4 kg/m^2^) for 24-hour ambulatory BP monitoring. We assessed nighttime BP and BP dipping (nighttime:daytime BP ratio). The area deprivation index (**ADI**) was used to measure neighborhood deprivation. Associations between ADI and ambulatory BP were examined.

**Results:** Black participants exhibited higher nighttime diastolic BP compared with White participants (63±8 mmHg vs 58±7 mmHg, *p*=0.003), and attenuated BP dipping ratios for both systolic (0.92±0.06 vs 0.86±0.05, *p*=0.001) and diastolic BP (0.86±0.09 vs 0.78±0.08, *p*=0.007). Black participants experienced greater neighborhood deprivation compared with White participants (ADI scores: 110±8 vs 97±21, *p*<0.001), and ADI was associated with attenuated systolic BP dipping (ρ=0.342, *p*=0.019).

**Conclusions:** Our findings suggest neighborhood deprivation may contribute to higher nighttime BP and attenuated BP dipping, which are prognostic of CVD, and more prevalent in Black adults. Targeted interventions to mitigate the effects of neighborhood deprivation may help to improve nighttime BP.

**Clinical Trial Registry:** URL: https://www.clinicaltrials.gov; Unique identifier: NCT04576338

**Clinical Perspective:**

- We demonstrate young Black adults exhibit higher nighttime blood pressure and attenuated nighttime blood pressure dipping compared with young White adults.
- Black adults were exposed to greater neighborhood deprivation, which was associated with attenuated blood pressure dipping, irrespective of race.
- Our findings add to a growing body of literature indicating neighborhood deprivation may contribute to increased cardiovascular disease risk.

## Introduction

Cardiovascular disease (**CVD**) is the leading cause of death worldwide and is responsible for approximately 875,000 deaths annually in the United States.^1^ High blood pressure (**BP**), or hypertension, is a major contributor to CVD.^1,2^ Black Americans exhibit the highest prevalence and severity of hypertension in the United States and develop elevated BP at an earlier age than other racial/ethnic groups.^3–5^ Accumulating evidence demonstrates social determinants of health (**SDoH**), such as neighborhood deprivation^6^ and socioeconomic status^7^ are associated with adverse CVD-related health outcomes.^8^ For example, disadvantaged neighborhoods are more likely to be afflicted by higher crime rates and violence,^9^ which may promote stress and unhealthy coping behaviors that may contribute to elevated BP.^10^ Compared with other racial/ethnic groups in the United States, Black Americans are disproportionately exposed to neighborhood deprivation. For example, predominately minority neighborhoods are more likely to have limited access to safe spaces for leisure-time physical activity and healthful foods.^11,12^ These neighborhood characteristics may influence health behaviors that are associated with BP such as physical activity, sleep, and diet.^13–15^ Collectively, these findings suggest that the higher prevalence of elevated BP in Black Americans may be influenced, in part, by neighborhood deprivation.

Ambulatory 24-hour BP monitoring provides a comprehensive assessment of BP through the acquisition of daytime and nighttime BP measures.^16^ Thus, ambulatory BP monitoring is an advantageous tool when confirming a diagnosis and identifying BP phenotypes that would not be detected from a single clinic BP measurement.^17^ Further, nighttime BP is superior to daytime BP in predicting CVD-related events in younger and older individuals with and without hypertension.^18,19^ Lack of nighttime BP dipping (i.e., non-dipping; ≤10% decrease in BP at night)^20^ is also associated with CVD morbidities such as heart failure, stroke, and end target organ damage.^21^ Black Americans exhibit an increased risk for elevated nighttime BP and attenuated nighttime BP dipping,^22^ a phenomenon that may begin at an early age.^23^ Our prior data indicate there are racial disparities in clinic BP in young adults,^24–26^ and neighborhood deprivation is a significant mediator.^24^ However, relations between SDoH (i.e., neighborhood deprivation) and health behaviors with ambulatory BP are understudied.

The primary aim of this investigation was to examine associations between neighborhood deprivation with ambulatory BP in young Black and White adults. We also assessed health behaviors including physical activity, sleep, and dietary intake. We hypothesized Black participants would exhibit higher nighttime BP and attenuated BP dipping compared with White participants. Additionally, we hypothesized greater neighborhood deprivation and poorer health behaviors would be associated with worse ambulatory BP.

An exploratory aim of this investigation was to determine whether sodium-regulatory pathways that play a critical role in BP differed by race and whether they were associated with ambulatory BP or neighborhood deprivation. Specifically, we assessed renin-angiotensin-aldosterone system (**RAAS**) hormones (i.e., serum aldosterone and plasma renin activity (**PRA**), and urinary endothelin-1 (**ET-1**). Prior data indicate serum aldosterone and PRA are lower in Black adults compared with White adults.^27^ This is important as suppressed basal RAAS activity may hinder the ability to regulate BP,^28^ particularly in the context of high dietary sodium, which is ubiquitous in the United States.^29^ Further, urinary ET-1 is a 21-amino acid natriuretic peptide and is critical for sodium balance and BP regulation.^30^ Previous work suggests adults with hypertension demonstrate elevated renal ET-1 production, reported as urinary ET-1 excretion, compared with adults with normal BP.^31,32^ However, whether these racial differences in RAAS and potentially urinary ET-1 excretion may influence racial differences in ambulatory BP patterns remains unclear. We hypothesized Black participants may demonstrate lower blood concentrations of RAAS hormones and higher urinary ET-1 excretion than White participants, and that these biomarkers may be associated with ambulatory BP and neighborhood deprivation.

## Methods

### Study Participants

All participants provided written and verbal consent prior to engaging in study activities. Study protocol and procedures were approved by the Institutional Review Board for Research Involving Human Subjects of Auburn University, and the trial was registered on clinicaltrials.gov (<u>NCT04576338</u>). Additional details of the parent study conducted from September 2018 to April 2019, have been described elsewhere.^15,24,26^ Participant ages ranged from 20-23 years old. Exclusion criteria included: individual history of cancer, diabetes, or any chronic disease; signs, or symptoms of disease (i.e., history of chest pain, dizziness, unusual shortness of breath, etc.); stage two hypertension (i.e., BP >140/90 mmHg);^2^ body mass index >35 kg/m^2^; or use of cardiovascular-acting medications. Female participants taking hormonal contraceptives (n=6) were permitted to participate, and the menstrual cycle was not controlled for during the study.

All participants reported that both of their parents were either ‘Black’ or ‘White’ and self-reported their race to be ‘Black’ or ‘White’. In addition to using self-report of race, a social construct, we also quantified skin pigmentation, which can influence physiology (e.g., vitamin D production)^33^ and social experiences (e.g., colorism).^34,35^ As previously described,^25,26,33^ skin pigmentation was measured as the melanin-index (M-index) using reflectance spectrophotometry (DSM3 DermaSpectrometer; Cortex Technology, Hadsund, Denmark). We measured the M-index on the inner aspect of the participant’s upper arm because this region is easily accessible and represents constitutive skin pigmentation due to its relatively low sun exposure.^33^

### Study Design

Residential information and SDoH questionnaires were collected in the parent study (September 2018 to April 2019).^15^ Health behaviors (i.e., physical activity, sleep, diet), BP measures (i.e., in-laboratory and ambulatory BP), and blood and urine samples were collected between January and December 2021.^26^ Detailed experimental procedures are described below, and an overview of the study design was as follows: after providing additional written consent, participants were provided actigraphy devices to characterize seven-day activity and sleep patterns, and a three-day food and fluid log for dietary assessment. Participants were outfitted with an ambulatory BP monitor and were provided with a urine collection container for simultaneous 24-hour urine collection prior to an in-person laboratory study visit where a venous blood sample was obtained.

### 24-hour Ambulatory Blood Pressure Monitoring

Participants wore a 24-hour ambulatory BP monitor (Suntech Oscar2) on their upper arm programmed to measure BP every 20 minutes during awake hours and every 30 minutes during asleep hours based on self-report by the participant.^17^ Participants were instructed to avoid alcohol, caffeine, and vigorous physical activity while wearing the monitor following their laboratory visit. Ambulatory BP data was extracted using AccuWin Pro software and processed by a single trained investigator (SJ). We applied the American Heart Association’s guidelines^17^ and defined daytime as 10 AM to 8 PM and nighttime as midnight to 6 AM.^36^ At least 20 daytime and 7 nighttime readings were required for a participant’s data to be included in the analysis.^17^ As previously described,^37^ we derived average daytime and nighttime BP, BP dip ratio (average nighttime BP divided by average daytime BP), and nighttime BP dipping (absolute difference between nighttime BP and daytime BP). BP dip ratios allow for the comparison of BP dipping in relation to daytime BP, thus providing a more comprehensive assessment of ambulatory BP patterns than absolute dipping alone.^38^

### Social Determinants of Health

#### Neighborhood socioeconomic disadvantage

Participants identified their residential address, or closest geographical unit (e.g., city, county) for each year of their life during their lab visit. To help identify and confirm the correct residential addresses, our data collection team worked with participants using Google Maps to confirm each address, and participants were encouraged to contact family members or friends to assist in recalling each address. Consistent with previous studies,^39^ we identified primary residential addresses for each of three developmental periods based on the number of years the participant lived in the neighborhood: early childhood (ages 0-5 years), middle childhood (6-12 years), and adolescence (13-18 years). The same residential address could be used as the primary residence for multiple developmental periods, if applicable.

We averaged the census tract level area deprivation index (**ADI**) scores across the three developmental periods to calculate a cumulative childhood ADI score. These Census survey years were chosen so that data for our sample, collected in 2018-2019 with an average age of 19 years at the time, closely corresponded to the three developmental periods.^15,24^ The ADI is a composite measure of neighborhood socioeconomic disadvantage composed of 17 poverty, education, housing, and employment census-tract indicators and has been previously used to track neighborhood-level disparities.^40^ Each indicator was independently weighed by factor scores to ensure poverty, income, and education had the largest relative weights. Similar to prior studies,^15,24^ the composite score was standardized to have a mean of 100 and a standard deviation (SD) of 20 to assist interpretation. Importantly, ADI is a validated approach for comprehensively assessing neighborhood deprivation.^41^ Details on additional SDoH variables measured in this study (i.e, socioeconomic status, perceived discrimination, and adverse childhood experiences) can be found in **Data S1-S3**.

### Health Behaviors

#### Physical activity and sleep

Participants were instructed to wear a validated tri-axial accelerometer^42^ (ActiGraph wGT3X-BT, Pensacola, FL, USA) on their right hip for a minimum of seven consecutive days with a minimum of seven days and 1,000 minutes of wear time per day prior to laboratory visit. Participants were instructed to wear an Actiwatch Spectrum Plus monitor (Philips Respironics) on the non-dominant wrist for 24 hours a day for seven to eight consecutive days and nights with a minimum of six nights prior to laboratory visit to be included in the analyses. Additional details regarding physical activity, sleep, and dietary assessment can be found in **Data S4-S6**.

### Urinary and Blood Metabolic Biomarkers

#### 24-hour urine collection

Urine was collected for 24 hours before the study visit in a light-protected, sterile 3.5 L container.^26,43^ Participants were instructed to record the time of their first void into a toilet on the day preceding the study visit, as well as their last void into the urine collection container on the day of the study visit. Urine flow rate was used to derive urinary ET-1 excretion and was calculated from urine volume and self-reported time the participant used the container in total. Based on National Health and Nutrition Examination Survey recommendations,^44^ we did not include data that required indexing to 24-hour urine flow rate for participants (n=3) who self-reported a urine collection time of fewer than 22 hours. We stored mixed aliquots from the 24-hour urine sample in cryogenic tubes at −80°C. *Blood and urine biochemical analysis* We assessed serum aldosterone and PRA to characterize potential racial differences in RAAS and examine associations with urinary ET-1 excretion and ambulatory BP. Serum aldosterone and PRA were measured in duplicate via radioimmunoassay at the Wake Forest University Biomarker Analytical Core. The inter-assay coefficient of variation was 6.8% for serum aldosterone and 5.3% for PRA. In addition, venous blood samples were obtained from vacutainers treated with dipotassium ethylenediaminetetraacetic acid. We assessed blood chemistry (i.e., glucose, lipids, and cholesterol) using an Alere Cholestech LDX analyzer, per manufacturer’s instructions (Abbott Laboratories, Abbott Park, IL, USA). We assessed urinary ET-1 excretion from stored urine samples as ET-1 elicits natriuretic effects and regulates BP. All samples were assayed in triplicate using a 96-well QuantiGlo ELISA kit (CN: QET00B, R&D Systems). The kit’s detection range was 0.023-0.102 pg/mL. The average intra-assay coefficient of variation for urinary ET-1 was 12.5%. We reported urinary ET-1 excretion as follows: concentration of the assayed sample (pg/mL) x urine flow rate (mL/min).

#### Statistical Analyses

Outliers were identified for primary outcomes including BP, ADI, and kidney variables using box plots. Any data points exceeding 1.5 times the length of the interquartile range (i.e., 1.5 times the range between the 1st and 3rd quartiles below the 1st and above the 3rd quartiles) were defined as outliers and removed from the dataset prior to analysis. We inspected variables for normality using the Shapiro-Wilk test and quantile-quantile plots. Racial comparisons were made using independent samples *t*-test for normally distributed data and the Mann-Whitney *U* test for non-normally distributed data. We used Cohen’s *d* to assess the effect size for normally distributed data,^45^ interpreted as small (*d* = 0.2-0.49), medium (*d* = 0.5-0.79), and large (*d* > 0.8). We used Rank-biserial correlation to assess effect size for non-normally distributed data,^46^ interpreted as small (0-0.19), medium (0.2-0.29), large (0.3-0.39), and very large (0.4-1.0). Ambulatory BP outcomes were compared by race and time (daytime and nighttime) using mixed-effect models. Sidak’s multiple comparisons were used to determine pairwise comparisons upon detecting significant main or interaction effects. Pearson’s *r* (normally distributed data) and Spearman’s *rho* (non-normally distributed data) correlations were conducted to assess associations between SDoH, health behaviors, and urinary ET-1 excretion with ambulatory BP measures. We used multiple regression models to examine outcomes associated with systolic BP and report standardized estimates and 95% confidence intervals for individual variables and adjusted R^2^ for overall model fit. Statistical analyses were completed using Jamovi 2.3.21.0 and GraphPad 9.5.0 (GraphPad Prism Software, San Diego, CA, USA) with *a priori* significance set as *p* ≤ 0.05 for all statistical analyses. Using American Psychological Association convention, when we report “*ps* ≤ 0.###” it represents the highest *p* value being discussed is less than 0.0### and vice versa for when we report “*ps* ≥ 0.###”.

## Results

Descriptive characteristics of our Black (n=21) and White (n=26) participants and blood and urine biomarkers are presented in **Table 1**. Black participants exhibited a higher M-index than White participants (*p* < 0.001). No racial differences in biological sex proportions, clinical BP values, or blood glucose or lipids were observed (*ps* ≥ 0.111). Black participants demonstrated lower serum aldosterone, but PRA was not different between Black and White participants. Lastly, we did not observe a racial difference in urinary ET-1 excretion (*p* = 0.592).

**Table 1.** Race comparison of participant characteristics and biochemical markers. Data reported in mean ± SD (normal distribution) or median (IQR) (non-normal distribution). Independent samples *t*-test was conducted for normally distributed data with Cohen’s *d* effect size. Mann-Whitney *U* test was conducted for non-normally distributed data with rank biserial correlation effect size. A Fisher’s exact test was conducted to compare distribution of biological sex between Black and White cohorts. *Significance was set to *p* ≤ 0.05 for all statistical analyses.

| Variables | Black participants<br>(n=21) | White participants<br>(n=26) | *p* value | Effect<br>size |
| --- | --- | --- | --- | --- |
| **Participant characteristics** |  |  |  |  |
| Age, years | 21 (1) | 21 (1) | 0.429 | 0.12 |
| Sex | 7 M / 14 F | 13 M / 13F | 0.260 | 0.17 |
| Height, m | 1.66 ± 0.08 | 1.73 ± 0.11 | **0.026\*** | - |
| Mass, kg | 72.2 ± 10.5 | 74.8 ± 13.4 | 0.485 | 0.21 |
| Body mass index, kg/m<sup>2</sup> | 26.1 ± 4.2 | 24.7 ± 3.7 | 0.225 | 0.36 |
| Systolic blood pressure, mmHg | 120 ± 11 | 117 ± 9 | 0.399 | 0.29 |
| Diastolic blood pressure, mmHg | 72 ± 11 | 68.0 ± 7 | 0.138 | 0.51 |
| Waist-to-hip ratio | 0.81 ± 0.04 | 0.82 ± 0.06 | 0.692 | 0.13 |
| Melanin index | 54 ± 10 | 28 ± 4 | **< 0.001\*** | 3.45 |
| **Blood biomarkers** |  |  |  |  |
| Glucose, mg/dl | 81 (10) | 87 (11) | 0.111 | 0.29 |
| Triglycerides, mg/dl | 46(20) | 62 (39) | 0.123 | 0.28 |
| Cholesterol, mg/dl | 153 ± 32 | 168 ± 30 | 0.138 | 0.48 |
| Low-density lipoprotein, mg/dl | 100 ± 27 | 95 ± 32 | 0.732 | 0.14 |
| High-density lipoprotein, mg/dl | 55 ± 15 | 56 ± 15 | 0.888 | 0.04 |
| Aldosterone, ng/dl | 9.25 (5.61) | 14.21 (11.63) | **0.017\*** | 0.44 |
| Plasma renin activity, ng/ml/hr | 0.50 (0.60) | 0.90 (1.05) | 0.118 | 0.29 |
| **Urinary biomarkers** |  |  |  |  |
| Endothelin-1 excretion, pg/min | 0.24 (0.26) | 0.27 (0.44) | 0.592 | 0.10 |

### Black participants exhibited elevated nighttime diastolic BP and attenuated BP dipping

Average daytime and nighttime systolic and diastolic BP split by race are presented in **Figure 1**. We observed a significant interaction (*p* = 0.003) and time effect (*p* < 0.001), but no main effect of race on ambulatory systolic BP (**Figure 1A**, *p* = 0.222). There were no significant post-hoc differences for ambulatory systolic BP. For diastolic BP, significant interaction (*p* = 0.021), time (*p* < 0.001), and race (*p* = 0.022) effects were observed (**Figure 1B**). Post-hoc testing revealed nighttime diastolic BP was higher in Black participants compared with White participants (*p* = 0.003). Black participants exhibited higher systolic (*p* = 0.001) and diastolic (*p* = 0.007) BP dip ratios (**Figure 1C**) and lower systolic (*p* = 0.008) and diastolic (*p* = 0.017) absolute BP dipping compared with White participants (**Figure 1D**).

**Figure 1.**
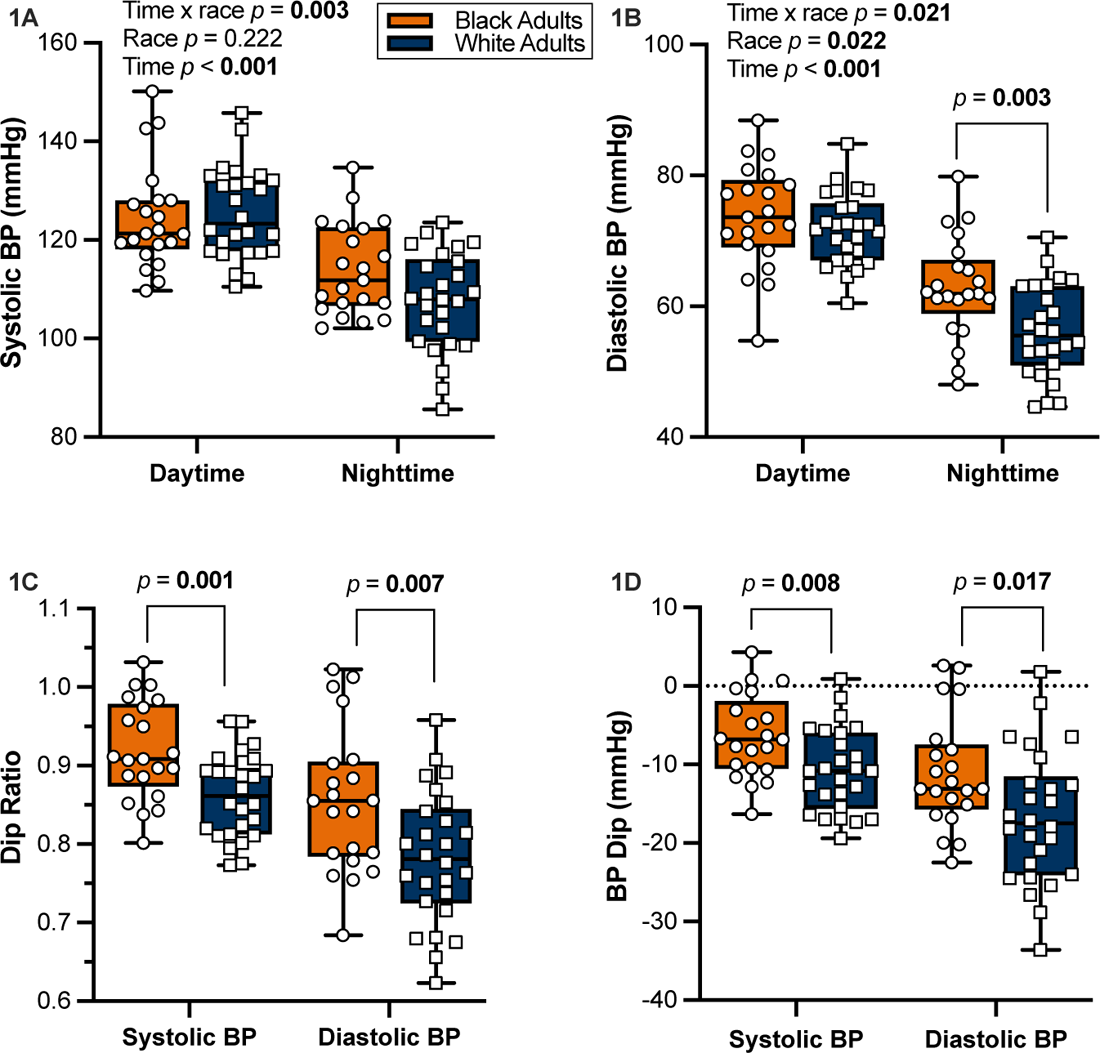
Race comparison of ambulatory blood pressure and dip ratios. Black participants (n=21) compared with White participants (n=26) for **(A)** daytime systolic BP (124 ± 9 mmHg vs 125 ± 11 mmHg) and nighttime (114 ± 9 mmHg vs 108 ± 10 mmHg); **(B)** daytime diastolic BP (74 ± 8 mmHg vs 72 ± 5 mmHg) and nighttime (63± 8 mmHg vs 58 ± 7 mmHg); **(C)** night-to-day systolic BP dip ratio (0.92 ± 0.06 vs 0.86 ± 0.05) and diastolic BP dip ratio (0.86 ± 0.09 vs 0.78 ± 0.08); and **(D)** absolute nighttime systolic BP dipping (6.4 ± 5.3 vs 10.7 ± 5.3 mmHg) and diastolic BP dipping (11.0 ± 7.2 vs 16.8 ± 8.5 mmHg). Mixed-effects model with Sidak’s multiple comparisons tests reported for A & B. Unpaired *t*-tests reported for C & D. Significance was set to *p* ≤ 0.05 for all statistical analyses.

### Race differences in SDoH and health behaviors and relations with ambulatory BP

Racial differences in SDoH and health behaviors are presented in **Table 2**. Black participants reported greater neighborhood deprivation than White participants for all developmental periods (*ps* ≤ 0.018). Black participants reported a lower income:needs ratio compared with White participants (*p* = 0.014) and Black participants reported higher ACEs than White participants (*p* = 0.028). In contrast, there was no racial difference in perceived discrimination (*p* = 0.329). Regarding health behaviors, Black participants reported fewer minutes of MVPA than White participants (*p* = 0.008). However, there was no racial difference in average daily steps or sedentary time (*ps* ≥ 0.067). Black participants also exhibited shorter sleep duration and poorer sleep efficiency than White participants (*ps* ≤ 0.024). Lastly, Black participants reported lower dietary total fiber, soluble fiber, insoluble fiber, calcium, magnesium, and potassium than White participants (*ps* ≤ 0.042).

**Table 2.**
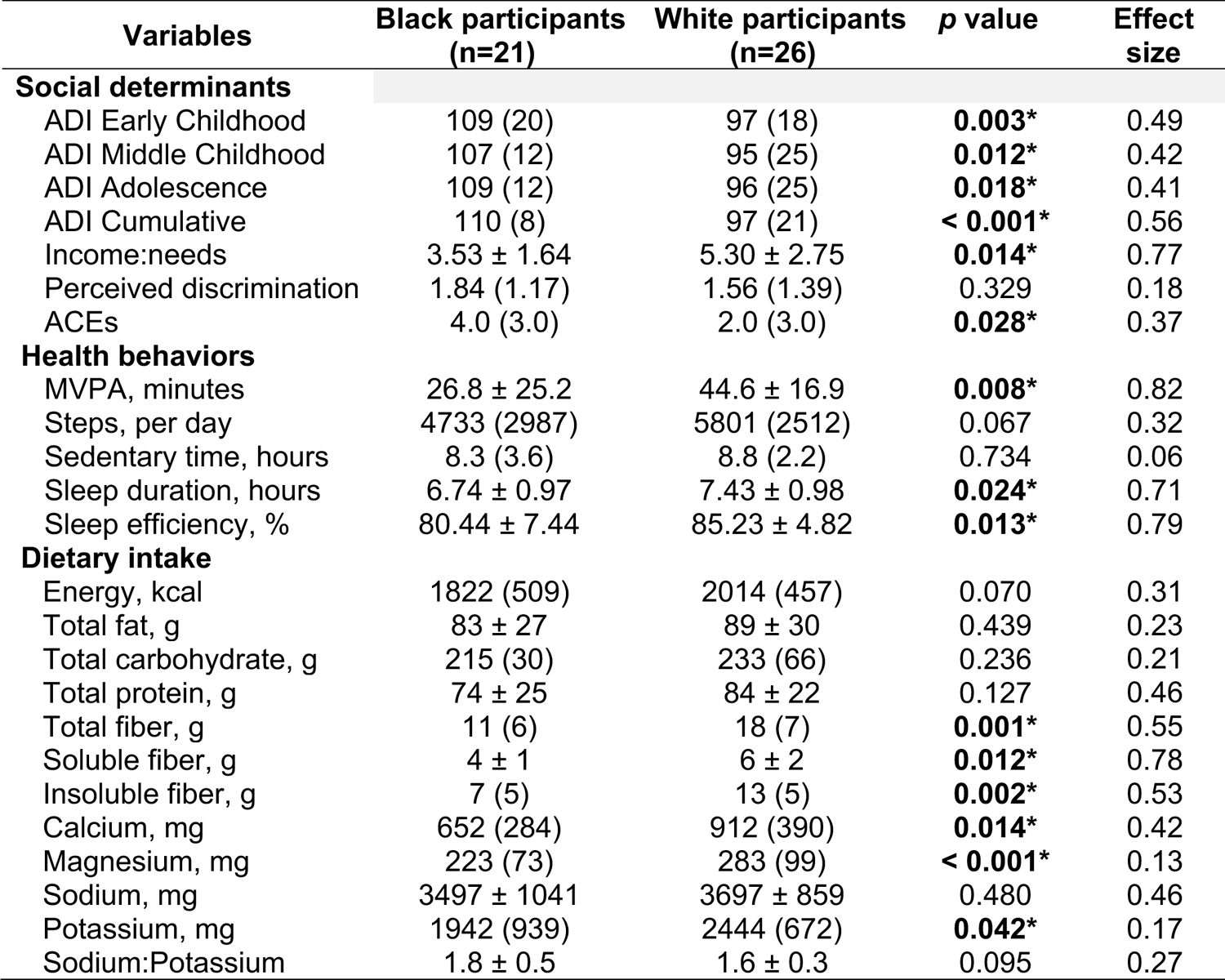
Race comparison of social determinants of health, health behaviors, and dietary intake. Data reported in mean ± SD (normal distribution) or median (IQR) (non-normal distribution). Independent samples *t*-test was conducted for normally distributed data with Cohen’s *d* effect size. Mann-Whitney *U* test was conducted for non-normally distributed data with rank biserial correlation effect size. Abbreviations include: area deprivation index (ADI), adverse childhood experiences (ACEs), and moderate to vigorous physical activity (MVPA). *Significance was set to *p* ≤ 0.05 for all statistical analyses.

| Variables | Black participants<br>(n=21) | White participants<br>(n=26) | *p* value | Effect<br>size |
| --- | --- | --- | --- | --- |
| **Social determinants** |  |  |  |  |
| ADI Early Childhood | 109 (20) | 97 (18) | **0.003\*** | 0.49 |
| ADI Middle Childhood | 107 (12) | 95 (25) | **0.012\*** | 0.42 |
| ADI Adolescence | 109 (12) | 96 (25) | **0.018\*** | 0.41 |
| ADI Cumulative | 110 (8) | 97 (21) | **< 0.001\*** | 0.56 |
| Income:needs | 3.53 ± 1.64 | 5.30 ± 2.75 | **0.014\*** | 0.77 |
| Perceived discrimination | 1.84 (1.17) | 1.56 (1.39) | 0.329 | 0.18 |
| ACEs | 4.0 (3.0) | 2.0 (3.0) | **0.028\*** | 0.37 |
| **Health behaviors** |  |  |  |  |
| MVPA, minutes | 26.8 ± 25.2 | 44.6 ± 16.9 | **0.008\*** | 0.82 |
| Steps, per day | 4733 (2987) | 5801 (2512) | 0.067 | 0.32 |
| Sedentary time, hours | 8.3 (3.6) | 8.8 (2.2) | 0.734 | 0.06 |
| Sleep duration, hours | 6.74 ± 0.97 | 7.43 ± 0.98 | **0.024\*** | 0.71 |
| Sleep efficiency, % | 80.44 ± 7.44 | 85.23 ± 4.82 | **0.013\*** | 0.79 |
| **Dietary intake** |  |  |  |  |
| Energy, kcal | 1822 (509) | 2014 (457) | 0.070 | 0.31 |
| Total fat, g | 83 ± 27 | 89 ± 30 | 0.439 | 0.23 |
| Total carbohydrate, g | 215 (30) | 233 (66) | 0.236 | 0.21 |
| Total protein, g | 74 ± 25 | 84 ± 22 | 0.127 | 0.46 |
| Total fiber, g | 11 (6) | 18 (7) | **0.001\*** | 0.55 |
| Soluble fiber, g | 4 ± 1 | 6 ± 2 | **0.012\*** | 0.78 |
| Insoluble fiber, g | 7 (5) | 13 (5) | **0.002\*** | 0.53 |
| Calcium, mg | 652 (284) | 912 (390) | **0.014\*** | 0.42 |
| Magnesium, mg | 223 (73) | 283 (99) | **< 0.001\*** | 0.13 |
| Sodium, mg | 3497 ± 1041 | 3697 ± 859 | 0.480 | 0.46 |
| Potassium, mg | 1942 (939) | 2444 (672) | **0.042\*** | 0.17 |
| Sodium:Potassium | 1.8 ± 0.5 | 1.6 ± 0.3 | 0.095 | 0.27 |

Associations between ADI and nighttime systolic BP are presented in **Figure 2A-D** and associations between ADI and systolic BP dip ratio are presented in **Figure 2E-H**. Corresponding associations between ADI and nighttime diastolic BP and diastolic BP dip ratio are presented in **Supplemental Figure 1**. Early childhood, middle childhood, adolescence, and cumulative ADI were not correlated with nighttime systolic (**Figure 2A-D**, *ps* ≥ 0.063) or diastolic BP (**Supplemental Figure 1A-D**, *ps* ≥ 0.160). Early childhood, middle childhood, and cumulative ADI were positively correlated with systolic BP dip ratio (**Figure 2E, F, H,** *ps* ≤ 0.044), but there were no associations between ADI and diastolic BP dip ratio (**Supplemental Figures 1E-H**, *ps* ≥ 0.314).

**Figure 2.**
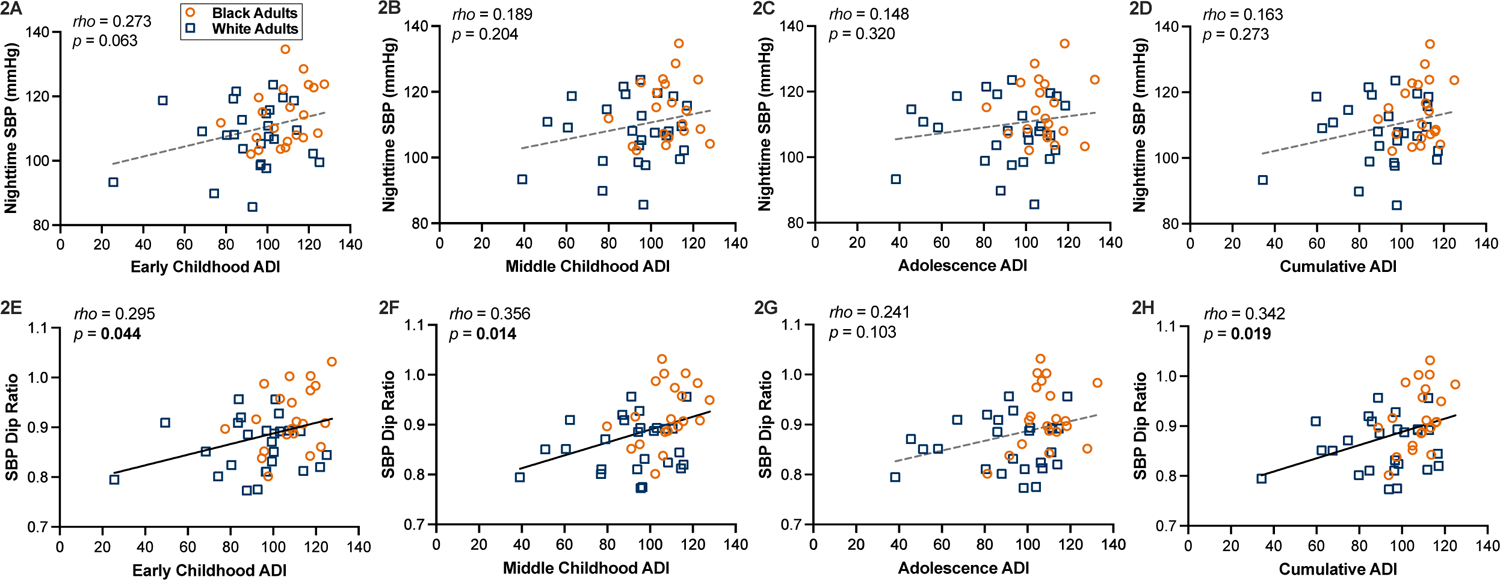
The associations between area deprivation index and nighttime systolic blood pressure. Spearman’s *rho* correlations in Black participants (n=21) and White participants (n=26) between nighttime systolic BP and ADI during **(A)** early childhood; **(B)** middle childhood; **(C)** adolescence; and **(D)** cumulative reported. Spearman’s *rho* correlations between systolic BP dip ratio and ADI during **(E)** early childhood; **(F)** middle childhood; **(G)** adolescence; and **(H)** cumulative reported. Abbreviations include: area deprivation index (ADI) and systolic blood pressure (SBP). Orange dots represent Black participants and blue squares represent White participants. Solid lines represent significant regressions. Significance was set to *p* ≤ 0.05 for all statistical analyses.

Additional associations between SDoH and health behaviors with nighttime systolic and diastolic BP, or systolic and diastolic BP dip ratios are presented in **Table 3**. Specifically, we performed bivariate correlations between the variables of interest with nighttime systolic and diastolic BP and systolic and diastolic BP dip ratios. Discrimination was positively correlated with nighttime diastolic BP (*p* = 0.044) but not nighttime systolic BP or systolic/diastolic BP dip ratios (*p*s ≥ 0.242). No other SDoH or health behavior was correlated with nighttime systolic and diastolic BP, nor systolic and diastolic BP dip ratios (*p*s ≥ 0.069).

**Table 3.**
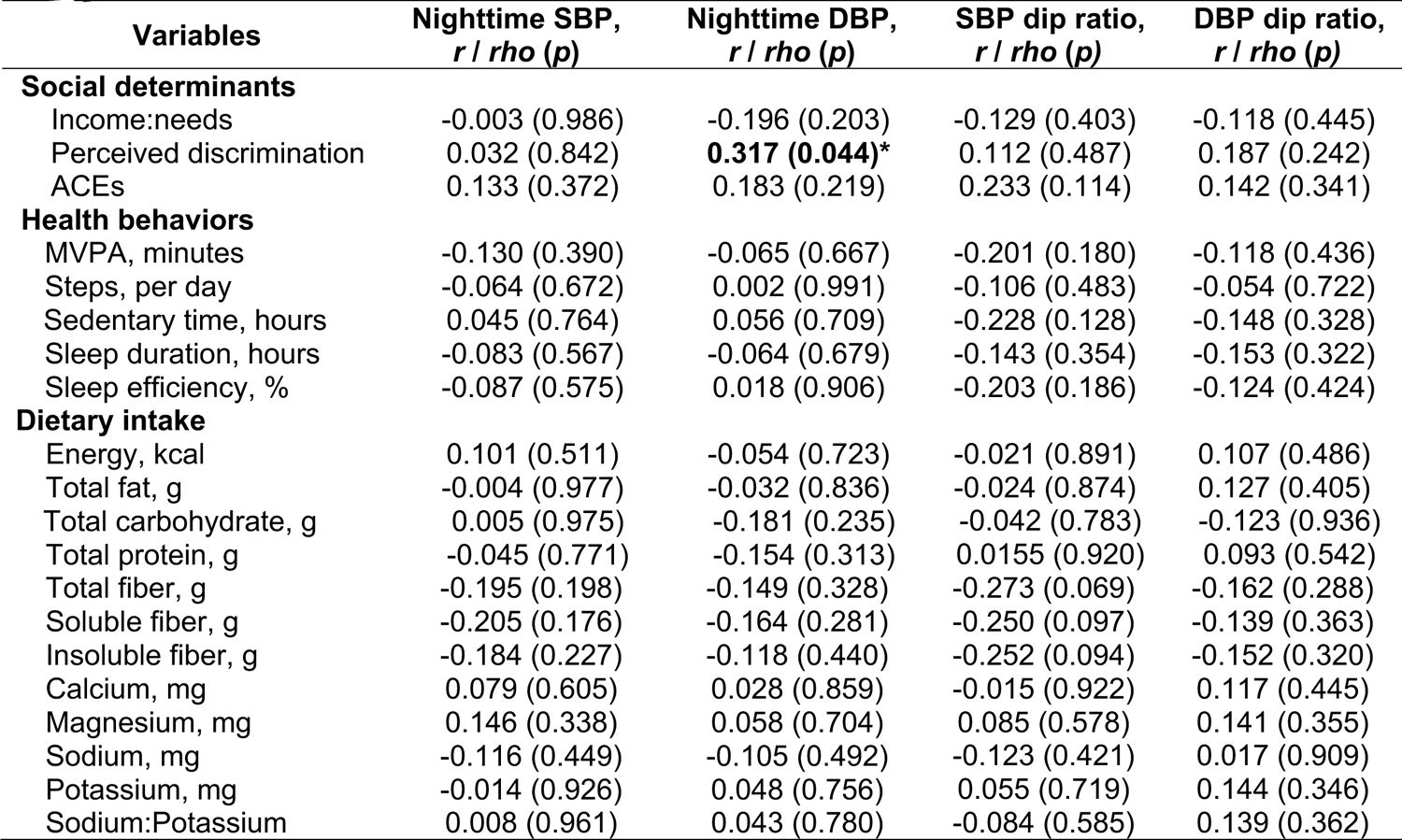
The associations between nighttime systolic and diastolic blood pressure and dip ratios with social determinants of health, health behaviors, and dietary intake. Pearson’s *r* and Spearman’s *rho* correlations reported (n=47). Abbreviations include: adverse childhood experiences (ACEs), diastolic blood pressure (DBP), moderate to vigorous physical activity (MVPA), and systolic blood pressure (SBP). *Significance was set to *p* ≤ 0.05 for all statistical analyses.

### Associations between sodium-regulatory biomarkers with ambulatory BP

Associations between aldosterone, PRA, and urinary ET-1 with nighttime systolic BP are presented in **Figure 3A-C** and associations between aldosterone, PRA, and urinary ET-1 and systolic BP dip ratio are presented in **Figure 3D-F**. Corresponding associations between aldosterone, PRA, and urinary ET-1 and nighttime diastolic BP and diastolic BP dip ratio are presented in **Supplemental Figure 2**. Aldosterone, PRA, and urinary ET-1 excretion were not correlated with nighttime systolic or diastolic BP (*ps* ≥ 0.844), nor systolic or diastolic BP dip ratios (*ps* ≥ 0.255). Associations between aldosterone, PRA, and urinary ET-1 with SDoH, health behaviors, and dietary intake are presented in **Supplemental Table 2**. Interestingly, PRA was correlated with sleep efficiency (*p* = 0.002) and magnesium (*p* = 0.027). There were no associations between urinary ET-1 excretion and SDoH (*ps* ≥ 0.174) or health behaviors (*ps* ≥ 0.109) except potassium intake was negatively correlated with urinary ET-1 excretion (*p* = 0.012).

**Figure 3.**
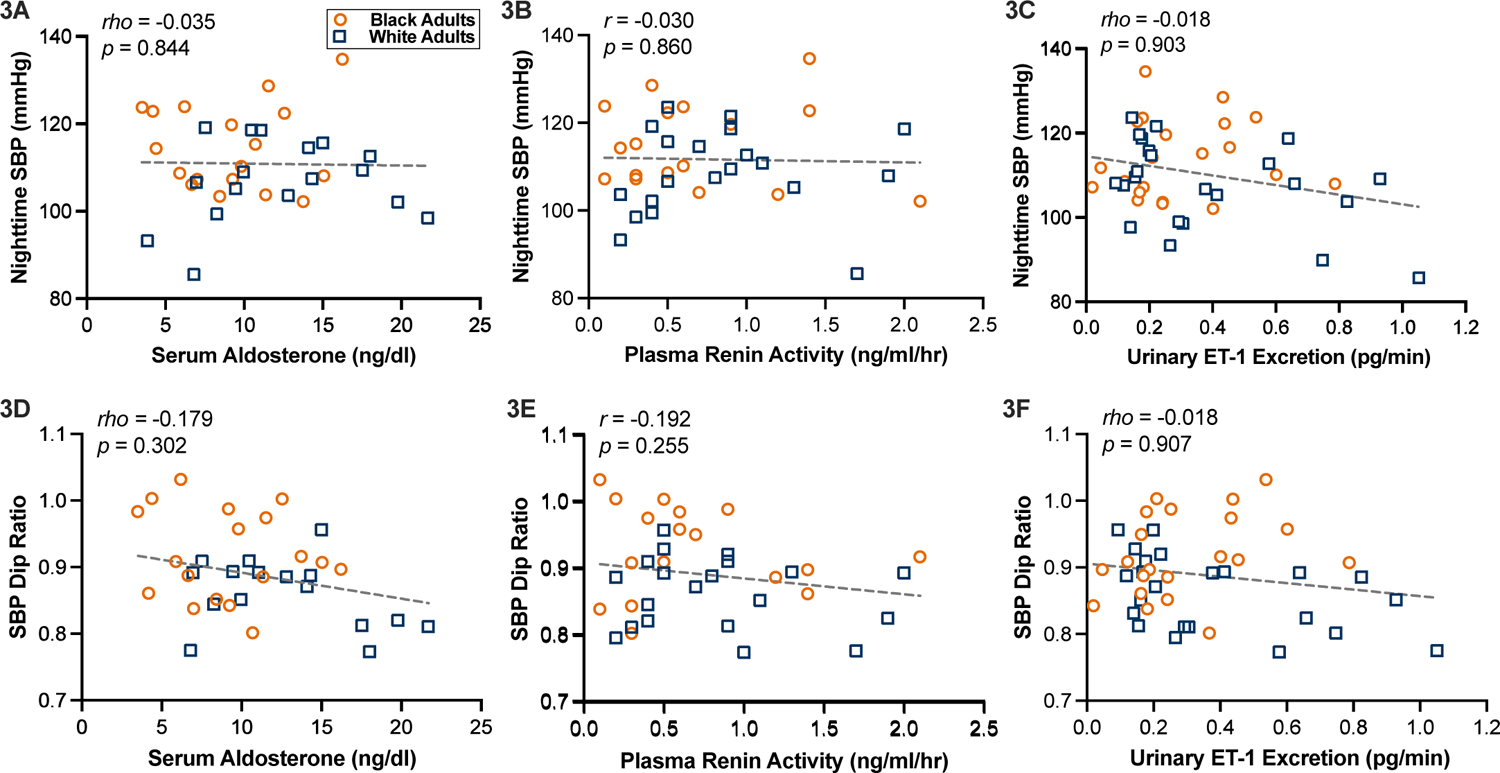
The associations between blood and urinary biomarkers and nighttime systolic blood pressure. Pearson’s *r* and Spearman’s *rho* correlations in Black participants (n≥17) and White participants (n≥17) between biomarkers with nighttime systolic BP and **(A)** serum aldosterone; **(B)** plasma renin activity; **(C)** urinary ET-1 excretion reported. Correlations between biomarkers with systolic BP dip ratio and **(D)** serum aldosterone; **(E)** plasma renin activity; **(F)** urinary ET-1 excretion reported. Abbreviations include: endothelin-1 (ET-1) and systolic blood pressure (SBP). Orange dots represent Black participants and blue squares represent White participants. 871 Significance was set to *p* ≤ 0.05 for all statistical analyses.

### Adjustment for social determinants of health and health behaviors on ambulatory BP

We performed a series multiple regressions to examine variables that may have potential independent associations with nighttime systolic BP, presented in **Table 4**. Race alone (model 1) was not associated with nighttime systolic BP (*β* = 0.545, *p* = 0.103). After controlling age, sex, and BMI (model 2), race (*β* = 0.876, *p* = 0.004) and sex (*β* = −0.599, *p* < 0.001) were associated with nighttime systolic BP. Further, after controlling for SDoH (model 3) and health behaviors (model 4), race (*β* = 1.180, *p* = 0.012) and sex (*β* = −0.747, *p* < 0.001) were independently associated with nighttime systolic BP and the overall model was significant (R^2^ = 0.341, *p* = 0.028). Similar multiple regression models were used to examine variables associated with systolic BP dip ratio (**Supplemental Table 3**) and nighttime diastolic BP and BP dip ratio (**Supplemental Tables 4 & 5**). In the fully adjusted model (model 4) for systolic BP dip ratio, race (*p* = 0.043), age (*p* = 0.042), and ACEs (*p* = 0.022) were associated with systolic BP dip ratio and the overall model was significant (R^2^ = 0.446, *p* = 0.006) (**Supplemental Table 3)**. In the fully adjusted model (model 4) for nighttime diastolic BP, race (*p* = 0.016) was associated with nighttime diastolic BP, but the overall model was not significant (**Supplemental Table 4)**. Regarding nighttime diastolic BP dip ratio, no individual variables were significantly associated, nor was the overall model significant (**Supplemental Table 5)**.

**Table 4.**
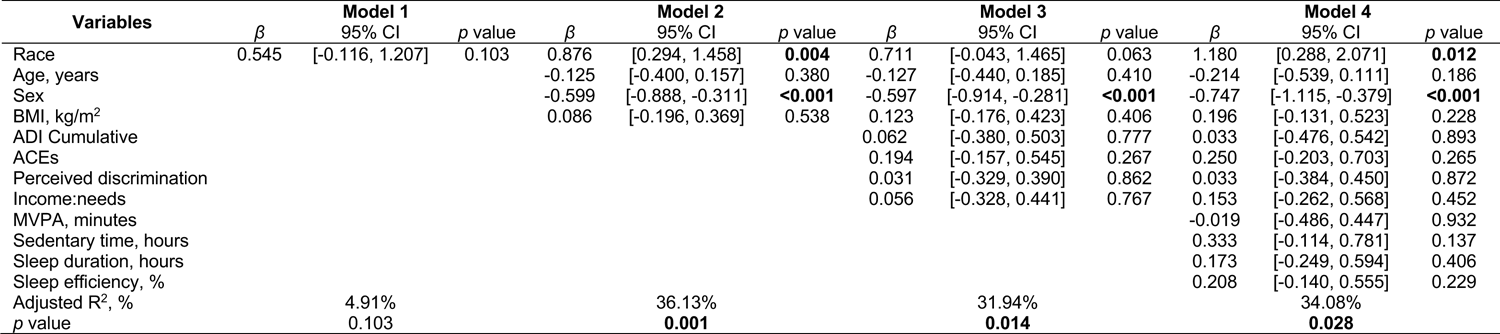
Multiple regression models on correlates of nighttime systolic BP. . Regression adjusted for race (**model 1**); adjusted for race, BMI, age, and sex (**model 2**), race, BMI, age, sex, and social determinants of health (**model 3**); race, BMI, age, sex, social determinants of health, and health behaviors (**model 4**). Standardized estimates (*β*) and 95% confidence intervals (95% CI) reported. Abbreviations include: body mass index (BMI), area deprivation index (ADI), adverse childhood experiences (ACEs), and moderate to vigorous physical activity (MVPA). Significance was set to *p_β_* ≤ 0.05 for all statistical analyses.

We performed an exploratory analysis comparing nighttime systolic BP dippers with non-dippers irrespective of race in **Supplemental Table 6**. Non-dippers reported greater ADI during middle childhood (*p* = 0.038) and exhibited lower MVPA compared to dippers (*p* = 0.029). Apart from these two variables there were no differences in SDoH, health behaviors, or blood/urine biomarkers in systolic BP dippers compared with non-dippers (*p*s ≥ 0.092). Lastly, we also performed preliminary sex comparisons, irrespective of race, for BP and blood/urine biomarkers which are reported in **Supplemental Table 7**. Compared with female participants, male participants exhibited higher screening systolic BP (*p* = 0.034) and nighttime systolic BP (*p* < 0.001). Male participants also exhibited higher PRA compared to female participants (*p* = 0.029). There were no sex differences for any of the remaining variables assessed (*p*s ≥ 0.252). Of the variables for which we observed a sex difference we explored potential main or interaction effects for sex and race. **Supplemental Figure 3** illustrates a race by sex comparison for nighttime systolic BP which revealed significant effects of race and sex (*p* < 0.001).

## Discussion

The purpose of this investigation was to examine associations between neighborhood deprivation and ambulatory BP in young Black and White adults. The main findings were that Black participants: 1) exhibited elevated nighttime diastolic BP and attenuated systolic and diastolic BP dipping; 2) experienced greater neighborhood deprivation, which was associated with attenuated systolic BP dipping; and 3) reported lower family income, more ACEs, and poorer scores on certain health behaviors (i.e., less MVPA and sleep, and worse dietary micronutrient intake) compared with White participants. Black participants also exhibited lower aldosterone than White participants in our study.

However, we did not observe racial differences in urinary ET-1 excretion and PRA, or an association between RAAS biomarkers or urinary ET-1 excretion and ambulatory BP. In contrast to our hypothesis, controlling for SDoH and health behaviors did not attenuate the association between race and nighttime BP. Nonetheless, this current investigation suggests notable racial differences in ambulatory blood pressure patterns, SDoH, and health behaviors among young adults. Together, these data suggest individuals who reside in, or have spent significant time residing in disadvantaged neighborhoods may be at higher risk for higher ambulatory BP, and consequently, increased CVD risk.

Consistent with our hypothesis and prior observations,^47,48^ we observed elevated nighttime diastolic BP and attenuated systolic and diastolic BP dipping (i.e., dip ratio and absolute dipping) in Black participants compared with White participants (**Figure 1**). Similar BP patterns were observed in a normotensive, healthy cohort of young adults (mean age: ∼22 years old) whereby Black adults had higher ambulatory diastolic BP than other racial/ethnic groups investigated.^49^ Alarmingly, racial differences in ambulatory BP have been observed in cohorts even younger than ours.^50,51^ Regarding BP dipping, there are data indicating Black individuals also exhibit a higher prevalence of non-dipping BP.^52,53^ Among adolescents, there are reports of Black adolescents exhibiting higher systolic BP dip ratio and an attenuated decrease in absolute BP compared with White adolescents, similar to the BP patterns observed in our group.^54^ Thus, our findings add to existing literature in demonstrating early signs of attenuated BP dipping in young Black Americans. Collectively, these findings emphasize the need for primary prevention and treatment strategies to control BP disparities in Black Americans, even in childhood prior to the development of hypertension (i.e., primordial prevention).^55^

We observed racial differences in neighborhood characteristics, specifically greater ADI in Black participants compared with White participants (**Table 2**). Importantly, neighborhood deprivation was correlated with systolic BP dip ratio (**Figure 2**). Interestingly, non-dippers reported higher middle childhood ADI (**Supplemental Table 6**). Our current findings are broadly in agreement with prior data from our group demonstrating neighborhood deprivation in early childhood was associated with diastolic BP, and explained 22% of the racial difference in BP between young Black and White adults.^24^ Regarding the higher ADI reported in Black compared with White participants in our investigation, the history of systemic racism in the United States has increased the likelihood of Black Americans facing substantial wealth inequality and live in segregated and disadvantaged neighborhoods.^24,56^ Prior work also suggests the role of neighborhood characteristics, segregation and safety, and historic redlining on cardiovascular health outcomes.^57,58,59^ Thus, our findings and prior literature suggest BP is influenced by environment and SDoH. In contrast to our hypothesis, racial differences in ambulatory BP (e.g., nighttime BP and BP dip ratios) were robust even after statistical adjustment for SDoH and health behaviors.

In regression models we generally did not observe independent associations between SDoH or health behaviors on our primary BP outcomes, and in many cases race remained associated with the BP outcome of interest. This finding may be attributed to unmeasured exposures within the domain of neighborhood, and potential latent effects that emerge later in life. For example, the association between some of the exposures we captured and physiologic dysregulation may not emerge until later ages (e.g., midlife) whereas the participants we studied were still young adults.^60^ Further, allostatic load from cumulative burden of psychosocial adversity influences numerous physiological systems, including BP regulation.^61^ Although our study captured some aspects of psychosocial adversity (i.e., ADI as a proxy for neighborhood exposures, ACEs, and perceived discrimination), we were not able to capture all aspects of subjective psychosocial adversity, inclusive of the cumulative effects of generational disadvantage, potential epigenetic modifications, heightened perceived stress, and other coping strategies that influence cardiovascular health. Nonetheless, our study highlights the influence of inequality including ADI, an objective measure of neighborhood, and discrimination on ambulatory BP which is prognostic of future cardiovascular events.^21^

Contrary to our findings, prior studies in young adults have demonstrated increased ACEs are associated with poorer CV health.^24,62^ The reasons for this discrepancy are unclear but may include the fact that we used a relatively affluent cohort of college students whereas other studies may have used community samples with a greater distribution of ACEs. While we did not observe a racial difference or association between perceived discrimination and BP dipping, perceived discrimination was positively correlated with nighttime systolic BP (**Table 3**). We acknowledge our finding of no racial difference in perceived discrimination contrasts with the prevailing notion that Black Americans experience higher perceived discrimination than White Americans. Possible reasons for this discrepancy include: 1) our participants were college students who may have experienced less discrimination than a community sample due to relative affluence and young age; and 2) our modest sample size which may have been underpowered to detect a difference.

We also observed racial differences in several health behaviors (i.e., MVPA and sleep), though relations between these measures and ambulatory BP were not observed. Regular physical activity is considered a ‘polypill’ for the prevention and treatment of numerous disease states and reduced CVD risk.^63^ However, we did not observe associations between physical activity (i.e., MVPA and steps) or sedentary time with ambulatory BP. Additionally, Black participants exhibited shorter sleep duration and poorer sleep efficiency than White participants, but we did not observe associations between sleep variables (i.e., duration and efficiency) and ambulatory BP. Consistent with prior literature on racial differences in nutritional intake from our team^25,26^ and others,^64^ Black participants reported lower dietary fiber, calcium, magnesium, and potassium compared with White participants (**Table 2**). Neighborhood environments impact availability and access to healthful food, and may contribute to racial differences in diet quality and ultimately heath disparities.^65^ Increased dietary fiber, calcium, magnesium, and potassium are associated with improved daytime and nighttime BP in adults with normal BP.^66^ However, we did not observe any association between diet and ambulatory BP in our group (**Table 3**). The lack of significant associations between health behaviors and BP is contrary to our hypothesis but may be explained by our relatively modest college-specific sample.

Our exploratory aim was to examine potential racial differences in urinary ET-1 excretion and its association with SDoH and ambulatory BP. Urinary ET-1 excretion is a reflection of renal ET-1 production and is associated with sodium excretion and regulating BP in the context of high dietary salt.^67^ In a healthy kidney, ET-1 plays a role in vasodilation while promoting vasoconstriction in a diseased kidney.^30^ Further, race-specific associations between ET-1 and sodium excretion (i.e., stronger in Black vs. White Americans) have been reported.^68^ Contrary to our hypothesis, we did not observe a racial difference in urinary ET-1 excretion (**Table 1**) or any associations between urinary ET-1 with ambulatory BP (**Figure 3 & Supplemental Figure 2**). Further, SDoH and health behaviors were generally not associated with urinary ET-1 excretion (**Supplemental Table 2**). A potential explanation for the lack of associations could be the timing of the urine collection. Urinary ET-1 exhibits diurnal variation, and is lowest during nighttime hours and is elevated during early daytime hours, suggesting diurnal patterns of urinary ET-1 excretion.^69^ However, we did not capture potential differences in circadian urinary ET-1 excretion from our single 24-hour urine samples which did allow us to assess potential diurnal differences. Regarding RAAS, in agreement with previous data,^27,70,71^ Black participants in the current study exhibited lower aldosterone. While PRA was not statistically different, it was directionally lower in Black participants with a medium effect size (Rank biserial correlation = 0.29). Future studies may consider investigating additional sodium regulatory pathways in addition to ET-1, aldosterone, and PRA that possess endogenous circadian rhythms with nighttime BP.

### Limitations

Our findings should be interpreted in the context of the following limitations. A potential critique of our study is we did not control for menstrual cycle or hormonal contraception. However, the argument has been made not controlling for menstrual cycle in studies not specifically focused on the effect of menstrual phase increases ecological validity.^72^ Importantly, we used identical approaches for both Black and White participants, and a prior investigation reported ambulatory BP is not appreciably different across the menstrual cycle.^73^ Another limitation is an *a priori* power analysis was not conducted for this small clinical trial (NCT04576338), as the goal was simply to recruit as many participants as possible from the larger parent study during COVID-19. Last, census tract measures of neighborhood socioeconomic disadvantage are crude proxies for a range of social and physical environment factors and may not reflect an individual’s lived experience. However, the ADI is a reliable and objective composite measure to track neighborhood-level disparities. Future studies focused on assessing perceptions of neighborhood characteristics and individual experiences will complement the findings from the present study. Strengths of this study include multiple measures of SDoH, comprehensively characterizing the health behaviors our participants (i.e., physical activity, sleep, diet) and 24-hour ambulatory BP.

## Conclusion

We observed elevated nighttime diastolic BP and attenuated BP dipping in young Black participants compared with young White participants. Black participants also reported greater neighborhood deprivation, which correlated with systolic BP dipping. Additionally, we observed less physical activity, shorter sleep duration and poorer sleep efficiency, and indicators of poorer dietary intake in Black participants than White participants, though none of these variables were associated with nighttime BP or BP dipping. Longitudinal and quasi experiential studies are needed to further elucidate links between neighborhood deprivation, BP, and CVD risk. Such studies will inform efforts to address racial/ethnic and socioeconomic inequities in CVD and related risk factors.

## Non-standard Abbreviations and Acronyms

ACE: adverse childhood experience

BP: blood pressure

CVD: cardiovascular disease

DBP: diastolic blood pressure

ET-1: endothelin-1

M-index: melanin-index

MVPA: moderate to vigorous physical activity

PRA: plasma renin activity

SDoH: social determinants of health

SBP: systolic blood pressure

## Data Availability

Data will be made available by the corresponding author upon reasonable request.

## Acknowledgments

The authors thank the individuals who participated in this study. We also thank the form Auburn University undergraduate students Catherine Garrett, Sydney Jones, Ben Loftis, Sarah Nix, and Julia Nunn for their assistance with data collection and analyses.

## Sources of Funding

This work was supported by National Institutes of Health (NIH) grants K01HL147998 and R15HL165325 (to ATR), R15HL140504 (to TEFR), UL1TR003096 (pilot funding to ATR and TL-1 Fellowships to SJ and BAL), K99/R00DK119413 (to EYG), Auburn University Presidential Graduate Research Fellowship (to BAL), and U.S. Department of Veterans Affairs Grants IK2RX003670 (to KB).

## Disclosures

None

## Conflict of Interest

None

## Author Contributions

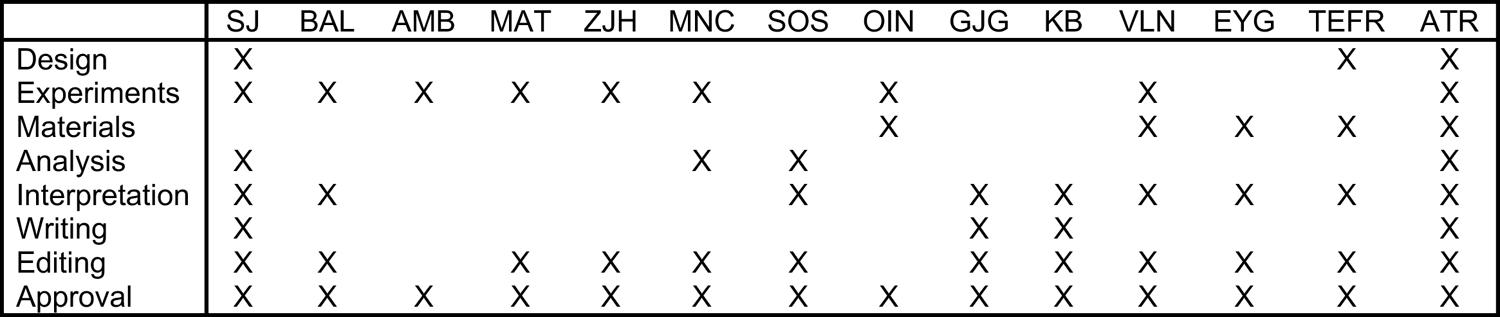

## Supplemental Data Appendix for

### Supplemental Methods

**Data S1. Socioeconomic status** The income-to-needs ratio, defined as family income divided by the poverty threshold for a given family size was used to index socioeconomic status. As previously described,^1^ household income was measured from participant reports of estimated parent income in the primary household on a scale of 1 (<$5,000) to 32 (>$500,000) with most intervals split by $5,000 increments (e.g., 16 = $75,000 to $79,999). The incomes were summed where applicable to create an estimated overall household income. Household income was divided by the corresponding U.S. Census poverty line based on family size to translate a family’s household income into an adjusted measure.

**Data S2. Perceived discrimination** To assess everyday discrimination, participants answered “The Everyday Discrimination Scale”, a series of survey questions regarding unfair treatment in the past year for nine specified scenarios (e.g., “you receive poorer service than other people at restaurants or stores” and “people act as if they are better than you”) with additional follow up questions focused on perceived reasons for those unfair treatments (e.g., your race, gender, etc.).^2^ Self-report responses were assigned a numeric scale and ranged from 1 (never) to 5 (once a week or more).^3^

**Data S3. Adverse childhood experiences** The number of adverse childhood experiences (**ACEs**) was measured using a 26-item inventory of different types of childhood adversity. Twelve of the items are based on the original and validated Adverse Childhood Experiences Questionnaire.^4^ Notably, several studies have called for addition of a more inclusive and robust number of potential ACEs to be considered that are associated with stress, particularly when examining socioeconomically and racially diverse populations.^5,6^ Therefore, we included the additional fourteen items which are based on childhood risk factors known to influence health including peer rejection and community violence exposure.^6^ Each item was measured by the occurrence (1 = yes, 0 = no), and summed to create a total score ranging from 0-26.

**Data S4. Physical activity** All devices were programmed using ActiLife software (ActiGraph Corp, Pensacola, FL, USA) and were set to record data at a sampling frequency of 30 Hz. Participants were also provided with an activity log to record their physical activity. On average, participants reported physical activity data for 8.1 ± 0.7 days. Participants were also provided with an activity log to document their physical activity and times the device was removed during daytime hours. Data were collected and analyzed in 60-second epochs. Wear time was assessed using the Choi algorithm^7^ and Montoye 2020 cut points were used to score physical activity.^8^ Steps per day were determined using ActiGraph’s default step algorithm. Physical activity intensity was scored as sedentary <2860 counts·min^−1^, light 2860-3940 counts·min^−1^, and moderate/vigorous (**MVPA**) were combined per developers’ recommendations and quantified as ≥3941 counts·min^−1^.^8^ Physical activity data scored in ActiLife was exported to Microsoft Excel to cross-check wear time with self-reported logs and to determine averages for step count and intensity.

**Data S5. Sleep** Philips Respironics devices use a solid-state piezo-electric sensor with a sampling rate of 32 Hz, and have been widely used and validated for sleep duration against polysomnography.^9^ On average, participants reported sleep actigraphy data for 7.5 ± 1.2 nights. Consistent with best practices,^10,11^ rest intervals were coded through visual inspection of the plotted data in Actiware software and using accelerations, self-reported markers, and daily sleep diary. If the monitor was removed, participants were asked to document the date and time of removal on their sleep diary. Sleep data were then visually inspected and manually adjusted with consideration of self-reported sleep logs by a single investigator (MNC). As previously described,^12,13^ actigraphy-derived sleep duration was operationalized as total nightly sleep time, not including wake after sleep onset, as time in bed and sleep duration are conceptually different outcomes. Sleep duration was quantified for each night the accelerometer was worn and averaged to determine participant mean values. Sleep efficiency was defined as the percent of time spent in bed the participant was truly asleep.^12^

**Data S6. Dietary assessment and analysis** Participants completed a three-day food and fluid record including at least two weekdays and one weekend day. To help with estimating portion sizes, participants were provided with a paper printout of two-dimensional food models.^14^ The diet record was returned at the laboratory testing visit, and a trained researcher (BAL and MAT) reviewed the record with the participant to obtain additional clarification. Diet records were analyzed for food group and nutrient intakes using the Nutrition Data System for Research (version 2020, University of Minnesota, Minneapolis, MN, USA) by a registered dietitian (SOS). Participants with at least two days of diet record data were included in the current analyses (mean = 3.0 ± 0.3 days). We reported the average nutrient intake for all reported days.

---

**Table S1.** The associations between area deprivation index and social determinants of health, health behaviors, and dietary intake. Pearson’s *r* and Spearman’s *rho* correlations reported. Abbreviations include: area deprivation index (ADI), adverse childhood experiences (ACEs), and moderate to vigorous physical activity (MVPA). *Significance was set to *p* ≤ 0.05 for all statistical analyses.

| ADI period | Early Childhood | Middle Childhood | Adolescence | Cumulative |
| --- | --- | --- | --- | --- |
| **Social determinants** |  |  |  |  |
| Income:needs | **-0.588 (<0.001)** | **-0.461 (0.002)** | **-0.353 (0.019)** | **-0.617 (<0.001)** |
| Perceived discrimination | 0.007 (0.966) | -0.133 (0.406) | 0.060 (0.713) | -0.069 (0.670) |
| ACEs | **0.334 (0.022)** | 0.220 (0.138) | 0.175 (0.240) | 0.263 (0.074) |
| **Health behaviors** |  |  |  |  |
| MVPA, minutes | -0.185 (0.218) | -0.289 (0.052) | -0.154 (0.307) | -0.218 (0.146) |
| Steps, per day | -0.209 (0.164) | -0.268 (0.072) | -0.086 (0.570) | -0.166 (0.268) |
| Sedentary time, hours | 0.094 (0.532) | 0.094 (0.535) | -0.038 (0.802) | -0.022 (0.885) |
| Sleep duration, hours | -0.106 (0.494) | -0.263 (0.084) | 0.087 (0.576) | -0.122 (0.431) |
| Sleep efficiency, % | 0.014 (0.926) | -0.014 (0.930) | 0.141 (0.363) | -0.045 (0.773) |
| **Dietary intake** |  |  |  |  |
| Energy, kcal | -0.094 (0.532) | 0.010 (0.943) | **-0.333 (0.024)** | -0.163 (0.280) |
| Total fat, g | -0.090 (0.549) | -0.069 (0.649) | **-0.400 (0.006)** | -0.196 (0.192) |
| Total carbohydrate, g | -0.113 (0.452) | 0.041 (0.787) | -0.217 (0.147) | -0.083 (0.582) |
| Total protein, g | -0.181 (0.227) | -0.035 (0.815) | **-0.339 (0.021)** | -0.189 (0.207) |
| Total fiber, g | **-0.386 (0.008)** | -0.248 (0.096) | **-0.341 (0.020)** | **-0.413 (0.005)** |
| Soluble fiber, g | -0.245 (0.101) | -0.107 (0.479) | -0.243 (0.104) | -0.244 (0.102) |
| Insoluble fiber, g | **-0.387 (0.008)** | -0.278 (0.062) | **-0.362 (0.013)** | **-0.434 (0.003)** |
| Calcium, mg | -0.038 (0.802) | 0.093 (0.537) | -0.169 (0.262) | -0.052 (0.733) |
| Magnesium, mg | -0.032 (0.834) | 0.181 (0.229) | -0.199 (0.186) | -0.004 (0.981) |
| Sodium, mg | -0.197 (0.188) | -0.057 (0.708) | **-0.342 (0.020)** | -0.227 (0.129) |
| Potassium, mg | 0.155 (0.301) | 0.072 (0.631) | -0.150 (0.320) | 0.104 (0.490) |
| Sodium:Potassium | 0.027 (0.859) | 0.031 (0.836) | -0.228 (0.127) | -0.047 (0.756) |

**Table S2.**
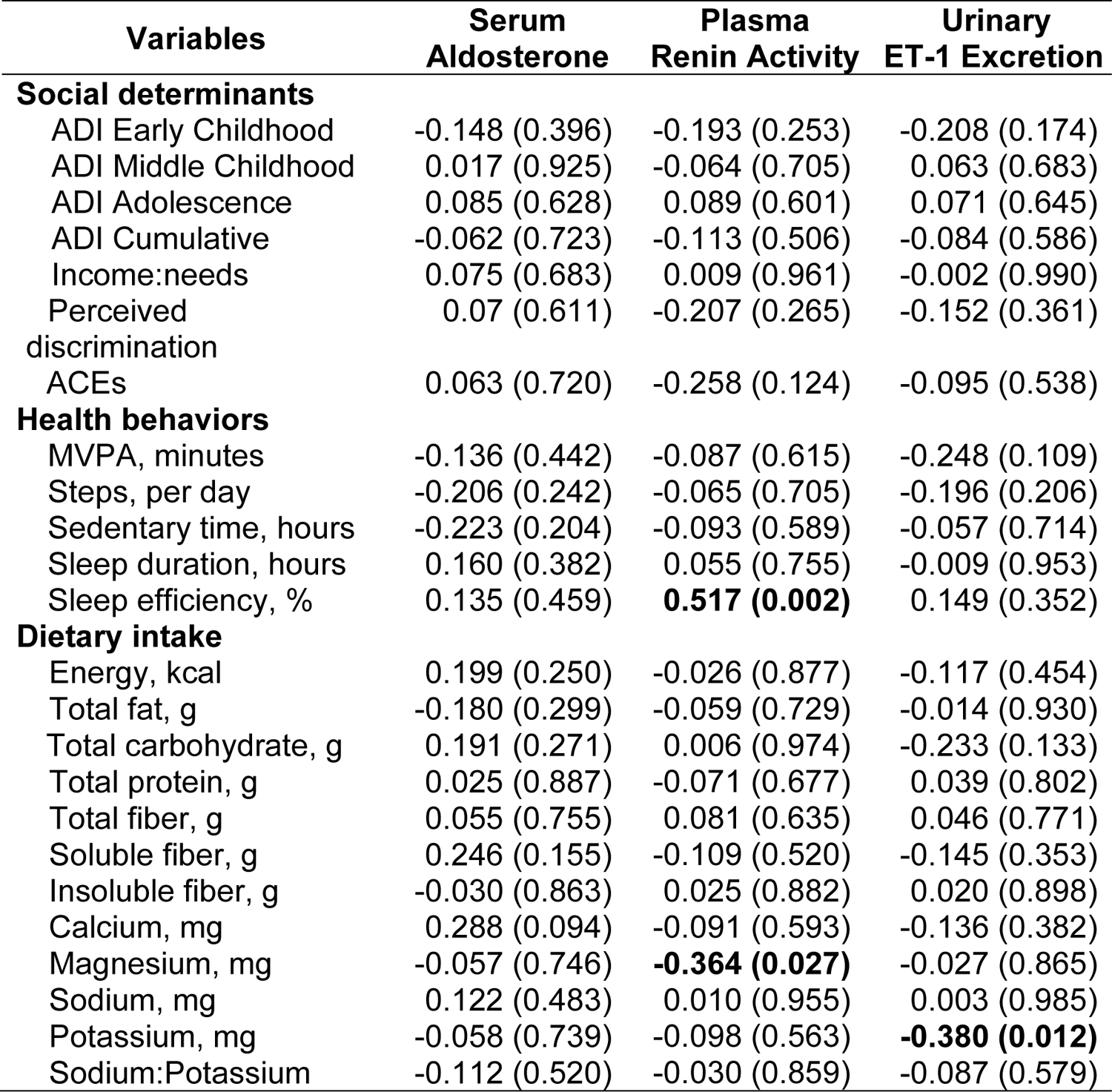
The associations between blood/urine biomarkers and social determinants of health, health behaviors, and dietary intake. Pearson’s *r* and Spearman’s *rho* correlations reported. Abbreviations include: area deprivation index 9ADI), adverse childhood experiences (ACEs), moderate to vigorous physical activity (MVPA), and endothelin-1 (ET-1). *Significance was set to *p* ≤ 0.05 for all statistical analyses.

**Table S3.** Multiple regression models on correlates of systolic BP dip ratio. Regression adjusted for race (**model 1**); adjusted for race, BMI, age, and sex (**model 2**), race, BMI, age, sex, and social determinants of health (**model 3**); race, BMI, age, sex, social determinants of health, and health behaviors (**model 4**). Standardized estimates (*β*) and 95% confidence intervals (95% CI) reported. Abbreviations include: body mass index (BMI), area deprivation index (ADI), adverse childhood experiences (ACEs), and moderate to vigorous physical activity (MVPA). Significance was set to *p* ≤ 0.05 for all statistical analyses.

| Variables | Model 1 |  |  | Model 2 |  |  | Model 3 |  |  | Model 4 |  |  |
| --- | --- | --- | --- | --- | --- | --- | --- | --- | --- | --- | --- | --- |
| | $\beta$ | 95% CI | *p* value | $\beta$ | 95% CI | *p* value | $\beta$ | 95% CI | *p* value | $\beta$ | 95% CI | *p* value |
| Race | 1.051 | [0.468, 1.634] | **<0.001** | 1.229 | [0.635, 1.822] | **<0.001** | 0.868 | [0.174, 1.563] | **0.016** | 0.846 | [0.028, 1.665] | **0.043** |
| Age, years |  |  |  | -0.241 | [-0.525, 0.043] | 0.093 | -0.287 | [-0.574, 0.001] | 0.051 | -0.310 | [-0.608, -0.012] | **0.042** |
| Sex |  |  |  | -0.258 | [-0.552, 0.036] | 0.084 | -0.212 | [-0.504, 0.080] | 0.147 | 0.062 | [-0.400, 0.276] | 0.707 |
| BMI, kg/m<sup>2</sup> |  |  |  | 0.060 | [-0.228, 0.348] | 0.676 | 0.113 | [-0.163, 0.389] | 0.409 | 0.196 | [-0.104, 0.497] | 0.190 |
| ADI Cumulative |  |  |  |  |  |  | 0.313 | [-0.094, 0.719] | 0.126 | 0.165 | [-0.303, 0.632] | 0.473 |
| ACEs |  |  |  |  |  |  | 0.272 | [-0.051, 0.596] | 0.096 | 0.495 | [0.079, 0.910] | **0.022** |
| Perceived discrimination |  |  |  |  |  |  | 0.060 | [-0.271, 0.391] | 0.713 | -0.147 | [-0.529, 0.236] | 0.436 |
| Income:needs |  |  |  |  |  |  | 0.285 | [-0.069, 0.639] | 0.111 | 0.244 | [-0.137, 0.625] | 0.198 |
| MVPA, minutes |  |  |  |  |  |  |  |  |  | 0.410 | [-0.018, 0.838] | 0.060 |
| Sedentary time, hours |  |  |  |  |  |  |  |  |  | -0.171 | [-0.581, 0.240] | 0.400 |
| Sleep duration, hours |  |  |  |  |  |  |  |  |  | -0.137 | [-0.524, 0.250] | 0.471 |
| Sleep efficiency, % |  |  |  |  |  |  |  |  |  | -0.248 | [-0.567, 0.071] | 0.122 |
| Adjusted R<sup>2</sup>, % |  | 26.20% |  |  | 33.69% |  |  | 42.23% |  |  | 44.46% |  |
| *p* value |  | **<0.001** |  |  | **0.002** |  |  | **0.002** |  |  | **0.006** |  |

**Table S4.** Multiple regression models on correlates of nighttime diastolic BP. Regression adjusted for race (**model 1**); adjusted for race, BMI, age, and sex (**model 2**), race, BMI, age, sex, and social determinants of health (**model 3**); race, BMI, age, sex, social determinants of health, and health behaviors (**model 4**). Standardized estimates (*β*) and 95% confidence intervals (95% CI) reported. Abbreviations include: body mass index (BMI), area deprivation index (ADI), adverse childhood experiences (ACEs), and moderate to vigorous physical activity (MVPA). Significance was set to *p* ≤ 0.05 for all statistical analyses.

| Variables | Model 1 |  |  | Model 2 |  |  | Model 3 |  |  | Model 4 |  |  |
| --- | --- | --- | --- | --- | --- | --- | --- | --- | --- | --- | --- | --- |
| | $\beta$ | 95% CI | *p* value | $\beta$ | 95% CI | *p* value | $\beta$ | 95% CI | *p* value | $\beta$ | 95% CI | *p* value |
| Race | 0.767 | [0.132, 1.401] | **0.019** | 0.879 | [0.208, 1.550] | **0.012** | 0.696 | [-0.176, 1.568] | 0.113 | 1.263 | [0.263, 2.263] | **0.016** |
| Age, years |  |  |  | -0.290 | [-0.611, 0.031] | 0.075 | -0.214 | [-0.576, 0.147] | 0.234 | -0.317 | [-0.681, 0.048] | 0.085 |
| Sex |  |  |  | 0.026 | [-0.307, 0.358] | 0.876 | 0.002 | [-0.365, 0.368] | 0.993 | -0.208 | [-0.621, 0.205] | 0.308 |
| BMI, kg/m<sup>2</sup> |  |  |  | -0.132 | [-0.457, 0.194] | 0.415 | -0.100 | [-0.447, 0.246] | 0.558 | 0.002 | [-0.365, 0.369] | 0.993 |
| ADI Cumulative |  |  |  |  |  |  | -0.007 | [-0.518, 0.504] | 0.977 | -0.061 | [-0.632, 0.510] | 0.827 |
| ACEs |  |  |  |  |  |  | 0.013 | [-0.394, 0.419] | 0.949 | 0.093 | [-0.415, 0.600] | 0.710 |
| Perceived discrimination |  |  |  |  |  |  | 0.240 | [-0.176, 0.656] | 0.246 | 0.246 | [-0.221, 0.714] | 0.287 |
| Income:needs |  |  |  |  |  |  | -0.031 | [-0.479, 0.414] | 0.888 | 0.094 | [-0.371, 0.559] | 0.680 |
| MVPA, minutes |  |  |  |  |  |  |  |  |  | -0.073 | [-0.596, 0.450] | 0.775 |
| Sedentary time, hours |  |  |  |  |  |  |  |  |  | 0.485 | [-0.017, 0.987] | 0.058 |
| Sleep duration, hours |  |  |  |  |  |  |  |  |  | 0.189 | [-0.284, 0.661] | 0.417 |
| Sleep efficiency, % |  |  |  |  |  |  |  |  |  | 0.255 | [-0.135, 0.645] | 0.190 |
| Adjusted R<sup>2</sup>, % |  | 12.57% |  |  | 15.20% |  |  | 8.87% |  |  | 17.06% |  |
| *p* value |  | **0.019** |  |  | 0.058 |  |  | 0.231 |  |  | 0.161 |  |

**Table S5.**
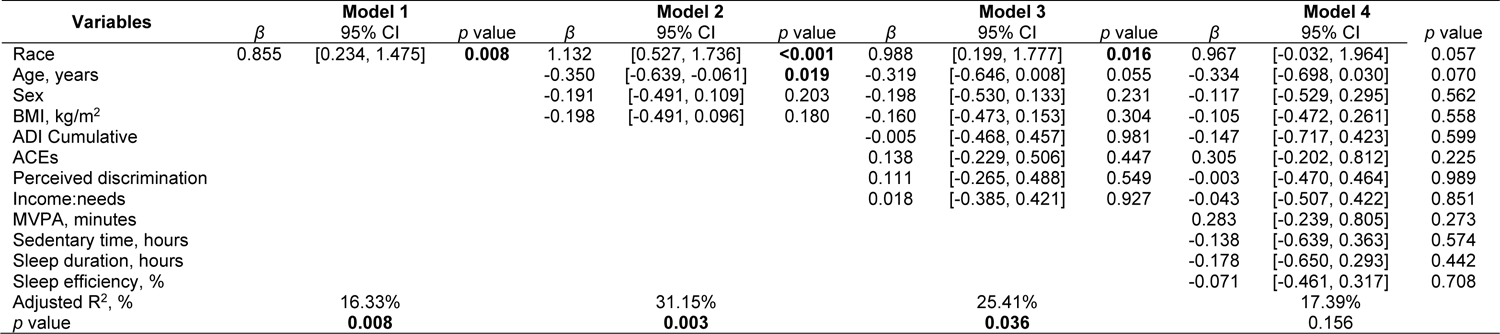
Multiple regression models on correlates of diastolic BP dip ratio. Regression adjusted for race (**model 1**); adjusted for race, BMI, age, and sex (**model 2**), race, BMI, age, sex, and social determinants of health (**model 3**); race, BMI, age, sex, social determinants of health, and health behaviors (**model 4**). Standardized estimates (*β*) and 95% confidence intervals (95% CI) reported. Abbreviations include: body mass index (BMI), area deprivation index (ADI), adverse childhood experiences (ACEs), and moderate to vigorous physical activity (MVPA). Significance was set to *p* ≤ 0.05 for all statistical analyses.

**Table S6.** Comparisons of social determinants of health, health behaviors, and blood/urine biomarkers based on blood pressure dipping status. Mann-Whitney *U*-test and rank biserial correlation effect sizes reported. Systolic blood pressure dip ratios were classified as dippers (< 0.90) and non-dippers (≥ 0.90). Abbreviations include: area deprivation index (ADI), adverse childhood experiences (ACEs), moderate to vigorous physical activity (MVPA), and endothelin-1 (ET-1). *Significance set to *p* ≤ 0.05.

| Variables | Dippers vs Non-Dippers, *p* value | Dippers vs Non-Dippers, Effect size |
| --- | --- | --- |
| **Social determinants** |  |  |
| ADI Early Childhood | 0.263 | 0.20 |
| ADI Middle Childhood | **0.038\*** | 0.36 |
| ADI Adolescence | 0.784 | 0.05 |
| ADI Cumulative | 0.118 | 0.28 |
| Income:needs | 0.467 | 0.13 |
| Perceived discrimination | 0.842 | 0.04 |
| ACEs | 0.200 | 0.22 |
| **Health behaviors** |  |  |
| MVPA, minutes | **0.029\*** | 0.39 |
| Steps, per day | 0.136 | 0.27 |
| Sedentary time, hours | 0.268 | 0.20 |
| Sleep duration, hours | 0.092 | 0.31 |
| Sleep efficiency, % | 0.213 | 0.23 |
| Energy, kcal | 0.391 | 0.16 |
| Total fat, g | 0.787 | 0.05 |
| Total carbohydrate, g | 0.652 | 0.08 |
| Total protein, g | 0.964 | 0.01 |
| Total fiber, g | 0.153 | 0.26 |
| Soluble fiber, g | 0.189 | 0.24 |
| Insoluble fiber, g | 0.213 | 0.23 |
| Calcium, mg | 0.893 | 0.03 |
| Magnesium, mg | 0.719 | 0.07 |
| Sodium, mg | 0.457 | 0.14 |
| Potassium, mg | 0.513 | 0.12 |
| Sodium:Potassium | 0.857 | 0.03 |
| **Biomarkers** |  |  |
| Glucose, mg/dL | 0.862 | 0.035 |
| Triglycerides, mg/dL | 0.205 | 0.235 |
| Cholesterol, mg/dL | 0.687 | 0.130 |
| Low-density lipoprotein, mg/dL | 0.882 | 0.063 |
| High-density lipoprotein, mg/dL | 0.939 | 0.025 |
| Aldosterone, ng/dl | 0.408 | 0.175 |
| Plasma renin activity, ng/ml/hr | 0.364 | 0.179 |
| ET-1 excretion, pg/min | 0.962 | 0.011 |

**Table S7.** Sex comparison on blood pressure and kidney variables. Data reported in mean ± SD (normal distribution) or median (IQR) (non-normal distribution). Independent samples *t*-test was conducted for normally distributed data with Cohen’s *d* effect size. Mann-Whitney *U* test was conducted for non-normally distributed data with rank biserial correlation effect size. Abbreviations include: blood pressure (BP) and endothelin-1 (ET-1). *Significance was set to *p* ≤ 0.05 for all statistical analyses.

| Variables | Male | Female | *p* value | Effect size |
| --- | --- | --- | --- | --- |
| Screening systolic BP, mmHg | 110 ± 8 | 104 ± 10 | **0.034** | 0.647 |
| Screening diastolic BP, mmHg | 64 (11) | 63 (8) | 0.804 | 0.044 |
| Nighttime systolic BP, mmHg | 117 ± 8 | 106 ± 9 | **<0.001** | 1.381 |
| Nighttime diastolic BP, mmHg | 59 ± 9 | 59 ± 8 | 0.795 | 0.077 |
| Systolic BP dip ratio | 0.90 ± 0.06 | 0.88 ± 0.06 | 0.339 | 0.285 |
| Diastolic BP dip ratio | 0.83 ± 0.10 | 0.81 ± 0.09 | 0.407 | 0.247 |
| Aldosterone, ng/dl | 11.85 ± 4.00 | 9.97 ± 4.91 | 0.252 | 0.408 |
| Plasma renin activity, ng/ml/hr | 0.90 (0.60) | 0.45 (0.45) | **0.029** | 0.424 |
| Urinary ET-1 excretion, pg/min | 0.20 (0.27) | 0.27 (0.28) | 0.573 | 0.103 |

## Supplemental Figures

**Figure S1.**
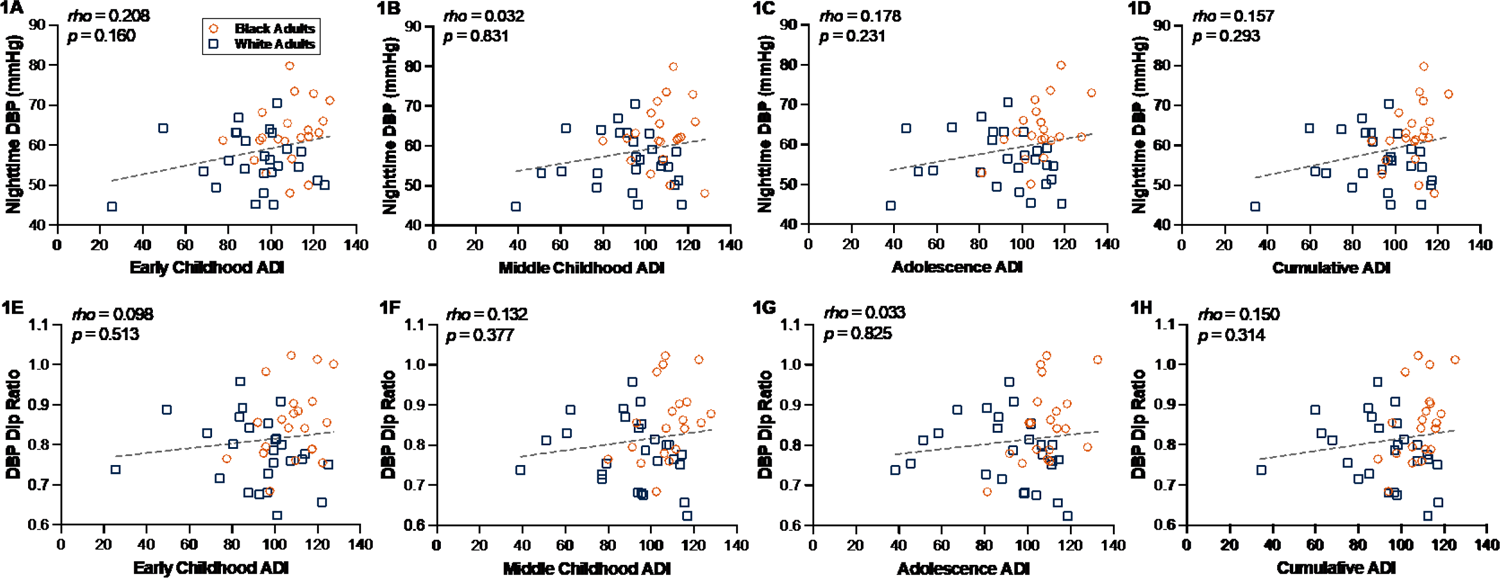
The associations between area deprivation index (ADI) and nighttime diastolic blood pressure. Spearman’s *rho* correlations in Black participants (n=21) and White participants (n=26) between nighttime diastolic BP and ADI during **(A)** early childhood; **(B)** middle childhood; **(C)** adolescence; and **(D)** cumulative reported. Spearman’s *rho* correlations between diastolic BP dip ratio and ADI during **(E)** early childhood; **(F)** middle childhood; **(G)** adolescence; and **(H)** cumulative reported. Orange dots represent Black participants and blue squares represent White participants. Significance was set to *p* ≤ 0.05 for all statistical analyses.

**Figure S2.**
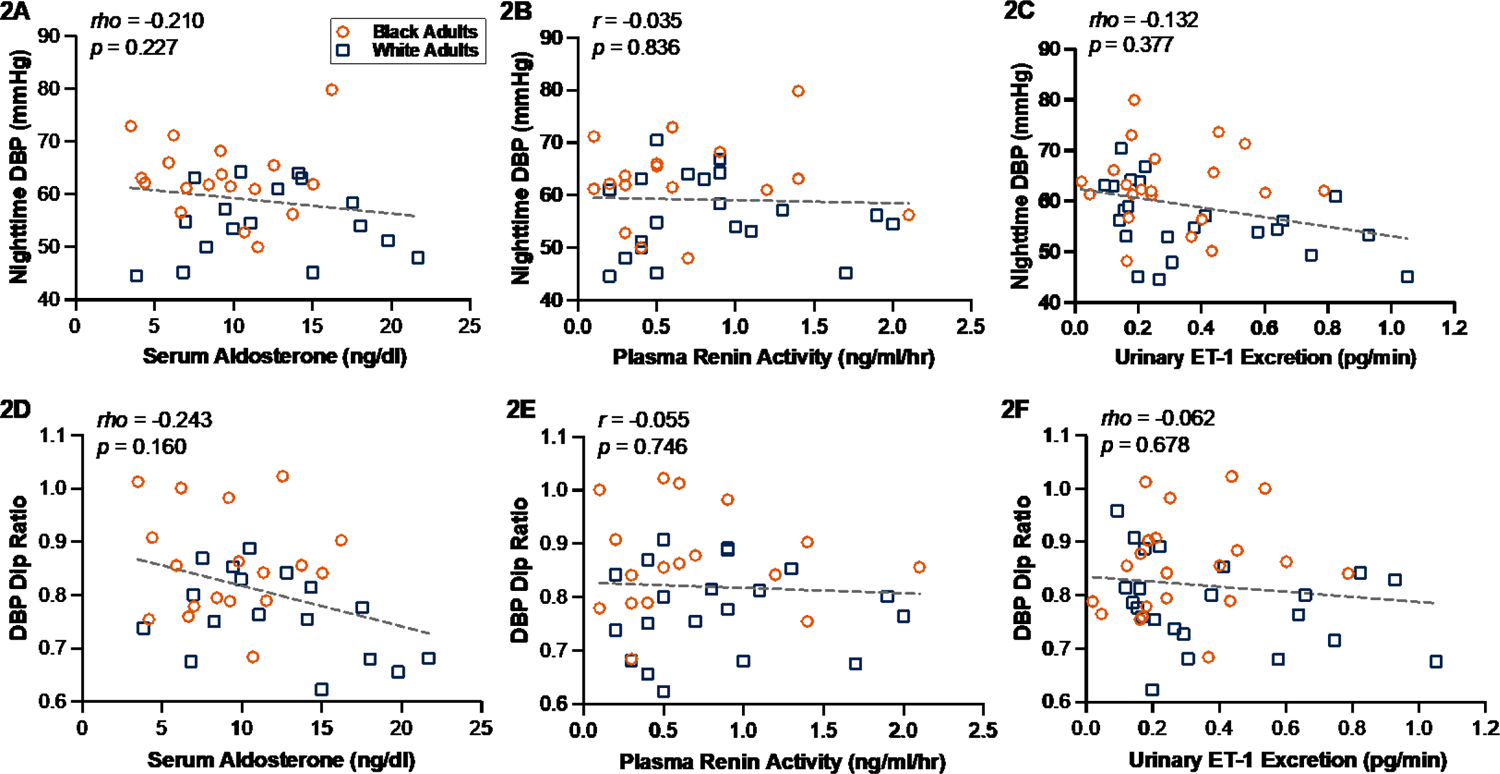
The associations between blood/urine biomarkers and nighttime diastolic blood pressure. Pearson’s *r* and Spearman’s *rho* correlations in Black participants (n≥17) and White participants (n≥17) between biomarkers with nighttime diastolic BP and **(A)** serum aldosterone; **(B)** plasma renin activity; **(C)** urinary ET-1 excretion reported. Correlations between biomarkers with diastolic BP dip ratio and **(D)** serum aldosterone; **(E)** plasma renin activity; **(F)** urinary ET-1 excretion reported. Abbreviations include: endothelin-1 (ET-1) and diastolic blood pressure (DBP). Orange dots represent Black participants and blue squares represent White participants. Significance was set to p ≤ 0.05 for all statistical analyses.

**Figure S3.**
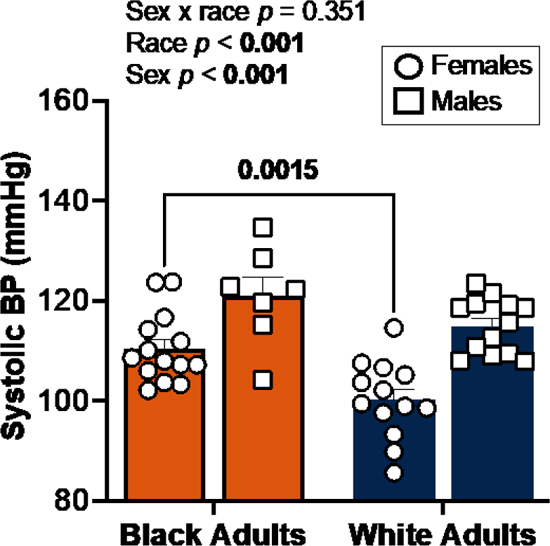
Race by sex comparison of nighttime systolic blood pressure. Data presented for Black males (mean: 121±10 mmHg), Black females (mean: 110±9 mmHg), White males (mean: 116±5 mmHg), and White females (mean: 100±8 mmHg). Orange bars represent Black participants and blue bars represent White participants. Circles represent females and squares represent males. Significance was set to *p* ≤ 0.05 for all statistical analyses.

## Notes

### Competing Interest Statement

The authors have declared no competing interest.

### Author Declarations

Study protocol and procedures were approved by the Institutional Review Board for Research Involving Human Subjects of Auburn University, and the trial was registered on clinicaltrials.gov (NCT04576338).

## References

1. Tsao CW, Aday AW, Almarzooq ZI, Anderson CAM, Arora P, Avery CL, Baker-Smith CM, Beaton AZ, Boehme AK, Buxton AE, et al. Heart Disease and Stroke Statistics-2023 Update: A Report From the American Heart Association. Circulation. 2023;147:e93–e621. doi: 10.1161/cir.0000000000001123

2. Fuchs FD, Whelton PK. High Blood Pressure and Cardiovascular Disease. Hypertension. 2020;75:285–292. doi: 10.1161/hypertensionaha.119.14240

3. Lackland DT. Racial differences in hypertension: implications for high blood pressure management. Am J Med Sci. 2014;348:135–138. doi: 10.1097/maj.0000000000000308

4. Carnethon MR, Pu J, Howard G, Albert MA, Anderson CAM, Bertoni AG, Mujahid MS, Palaniappan L, Taylor HA, Jr., Willis M, et al. Cardiovascular Health in African Americans: A Scientific Statement From the American Heart Association. Circulation. 2017;136:e393–e423. doi: 10.1161/cir.0000000000000534

5. Thomas SJ, Booth JN, Dai C, Li X, Allen N, Calhoun D, Carson AP, Gidding S, Lewis CE, Shikany JM, et al. Cumulative Incidence of Hypertension by 55 Years of Age in Blacks and Whites: The CARDIA Study. Journal of the American Heart Association. 2018;7:e007988. doi: 10.1161/jaha.117.007988

6. Barber S, Hickson DA, Kawachi I, Subramanian SV, Earls F. Neighborhood Disadvantage and Cumulative Biological Risk Among a Socioeconomically Diverse Sample of African American Adults: An Examination in the Jackson Heart Study. J Racial Ethn Health Disparities. 2016;3:444–456. doi: 10.1007/s40615-015-0157-0

7. Pickett KE, Pearl M. Multilevel analyses of neighbourhood socioeconomic context and health outcomes: a critical review. J Epidemiol Community Health. 2001;55:111–122. doi: 10.1136/jech.55.2.111

8. Grosicki GJ, Bunsawat K, Jeong S, Robinson AT. Racial and ethnic disparities in cardiometabolic disease and COVID-19 outcomes in White, Black/African American, and Latinx populations: Social determinants of health. Prog Cardiovasc Dis. 2022;71:4–10. doi: 10.1016/j.pcad.2022.04.004

9. Sampson RJ, Raudenbush SW, Earls F. Neighborhoods and violent crime: a multilevel study of collective efficacy. Science. 1997;277:918–924. doi: 10.1126/science.277.5328.918

10. Kuipers MA, van Poppel MN, van den Brink W, Wingen M, Kunst AE. The association between neighborhood disorder, social cohesion and hazardous alcohol use: a national multilevel study. Drug Alcohol Depend. 2012;126:27–34. doi: 10.1016/j.drugalcdep.2012.04.008

11. Jones SA, Moore LV, Moore K, Zagorski M, Brines SJ, Diez Roux AV, Evenson KR. Disparities in physical activity resource availability in six US regions. Preventive Medicine. 2015;78:17–22. doi: 10.1016/j.ypmed.2015.05.028

12. Bower KM, Thorpe RJ, Rohde C, Gaskin DJ. The intersection of neighborhood racial segregation, poverty, and urbanicity and its impact on food store availability in the United States. Preventive Medicine. 2014;58:33–39. doi: 10.1016/j.ypmed.2013.10.010

13. Barone Gibbs B, Hivert M-F, Jerome GJ, Kraus WE, Rosenkranz SK, Schorr EN, Spartano NL, Lobelo F. Physical Activity as a Critical Component of First-Line Treatment for Elevated Blood Pressure or Cholesterol: Who, What, and How?: A Scientific Statement From the American Heart Association. Hypertension. 2021;78. doi: 10.1161/hyp.0000000000000196

14. Ndanuko RN, Tapsell LC, Charlton KE, Neale EP, Batterham MJ. Dietary Patterns and Blood Pressure in Adults: A Systematic Review and Meta-Analysis of Randomized Controlled Trials. Adv Nutr. 2016;7:76–89. doi: 10.3945/an.115.009753

15. Fuller-Rowell TE, Nichols OI, Robinson AT, Boylan JM, Chae DH, El-Sheikh M. Racial disparities in sleep health between Black and White young adults: The role of neighborhood safety in childhood. Sleep Med. 2021;81:341–349. doi: 10.1016/j.sleep.2021.03.007

16. O’Brien E. Twenty-four-hour ambulatory blood pressure measurement in clinical practice and research: a critical review of a technique in need of implementation. Journal of Internal Medicine. 2011;269:478–495. doi: 10.1111/j.1365-2796.2011.02356.x

17. Muntner P, Shimbo D, Carey RM, Charleston JB, Gaillard T, Misra S, Myers MG, Ogedegbe G, Schwartz JE, Townsend RR, et al. Measurement of Blood Pressure in Humans: A Scientific Statement From the American Heart Association. Hypertension. 2019;73. doi: 10.1161/hyp.0000000000000087

18. Ohkubo T, Hozawa A, Yamaguchi J, Kikuya M, Ohmori K, Michimata M, Matsubara M, Hashimoto J, Hoshi H, Araki T, et al. Prognostic significance of the nocturnal decline in blood pressure in individuals with and without high 24-h blood pressure: the Ohasama study. J Hypertens. 2002;20:2183–2189. doi: 10.1097/00004872-200211000-00017

19. Staplin N, De La Sierra A, Ruilope LM, Emberson JR, Vinyoles E, Gorostidi M, Ruiz-Hurtado G, Segura J, Baigent C, Williams B. Relationship between clinic and ambulatory blood pressure and mortality: an observational cohort study in 59 124 patients. The Lancet. 2023. doi: 10.1016/s0140-6736(23)00733-x

20. Krzych ŁJ, Bochenek A. Blood pressure variability: Epidemiological and clinical issues. Cardiology Journal. 2013;20. doi: 10.5603/cj.2013.0022

21. Staessen JA. Predicting Cardiovascular Risk Using Conventional vs Ambulatory Blood Pressure in Older Patients With Systolic Hypertension. JAMA. 1999;282:539. doi: 10.1001/jama.282.6.539

22. Booth JN, Diaz KM, Seals SR, Sims M, Ravenell J, Muntner P, Shimbo D. Masked Hypertension and Cardiovascular Disease Events in a Prospective Cohort of Blacks. Hypertension. 2016;68:501–510. doi: 10.1161/hypertensionaha.116.07553

23. Hanevold CD. Racial-ethnic disparities in childhood hypertension. Pediatric Nephrology. 2023;38:619–623. doi: 10.1007/s00467-022-05707-x

24. Nichols OI, Fuller-Rowell TE, Robinson AT, Eugene D, Homandberg LK. Neighborhood Socioeconomic Deprivation in Early Childhood Mediates Racial Disparities in Blood Pressure in a College Student Sample. Journal of Youth and Adolescence. 2022;51:2146–2160. doi: 10.1007/s10964-022-01658-6

25. Grosicki GJ, Flatt AA, Cross BL, Vondrasek JD, Blumenburg WT, Lincoln ZR, Chall A, Bryan A, Patel RP, Ricart K, et al. Acute beetroot juice reduces blood pressure in young Black and White males but not females. Redox Biol. 2023;63:102718. doi: 10.1016/j.redox.2023.102718

26. Robinson AT, Linder BA, Barnett AM, Jeong S, Sanchez SO, Nichols OI, McIntosh MC, Hutchison ZJ, Tharpe MA, Watso JC, et al. Cross-sectional analysis of racial differences in hydration and neighborhood deprivation in young adults. Am J Clin Nutr. 2023. doi: 10.1016/j.ajcnut.2023.08.005

27. Wenner MM, Paul EP, Robinson AT, Rose WC, Farquhar WB. Acute NaCl Loading Reveals a Higher Blood Pressure for a Given Serum Sodium Level in African American Compared to Caucasian Adults. Front Physiol. 2018;9:1354. doi: 10.3389/fphys.2018.01354

28. Atlas SA. The Renin-Angiotensin Aldosterone System: Pathophysiological Role and Pharmacologic Inhibition. Journal of Managed Care Pharmacy. 2007;13:9–20. doi: 10.18553/jmcp.2007.13.s8-b.9

29. Hu JR, Sahni S, Mukamal KJ, Millar CL, Wu Y, Appel LJ, Juraschek SP. Dietary Sodium Intake and Sodium Density in the United States: Estimates From NHANES 2005-2006 and 2015-2016. Am J Hypertens. 2020;33:825–830. doi: 10.1093/ajh/hpaa104

30. Dhaun N, Goddard J, Kohan DE, Pollock DM, Schiffrin EL, Webb DJ. Role of Endothelin-1 in Clinical Hypertension. Hypertension. 2008;52:452–459. doi: 10.1161/hypertensionaha.108.117366

31. Mathur S, Pollock JS, Mathur S, Harshfield GA, Pollock DM. Relation of urinary endothelin-1 to stress-induced pressure natriuresis in healthy adolescents. Journal of the American Society of Hypertension. 2018;12:34–41. doi: 10.1016/j.jash.2017.11.005

32. Boesen EI. Endothelin receptors, renal effects and blood pressure. Current Opinion in Pharmacology. 2015;21:25–34. doi: 10.1016/j.coph.2014.12.007

33. Wolf ST, Jablonski NG, Ferguson SB, Alexander LM, Kenney WL. Four weeks of vitamin D supplementation improves nitric oxide-mediated microvascular function in college-aged African Americans. Am J Physiol Heart Circ Physiol. 2020;319:H906–h914. doi: 10.1152/ajpheart.00631.2020

34. Nguyen AW, Taylor HO, Keith VM, Qin W, Mitchell UA. Discrimination and social isolation among African Americans: The moderating role of skin tone. Cultur Divers Ethnic Minor Psychol. 2022. doi: 10.1037/cdp0000569

35. Burch T. Skin color and the criminal justice system: Beyond black-white disparities in sentencing. Journal of Empirical Legal Studies. 2015;12:395–420.

36. Bromfield SG, Booth JN, Loop MS, Schwartz JE, Seals SR, Thomas SJ, Min Y-I, Ogedegbe G, Shimbo D, Muntner P. Evaluating different criteria for defining a complete ambulatory blood pressure monitoring recording. Blood Pressure Monitoring. 2018;23:103–111. doi: 10.1097/mbp.0000000000000309

37. Migdal KU, Babcock MC, Robinson AT, Watso JC, Wenner MM, Stocker SD, Farquhar WB. The Impact of High Dietary Sodium Consumption on Blood Pressure Variability in Healthy, Young Adults. Am J Hypertens. 2020. doi: 10.1093/ajh/hpaa014

38. Fagard RH, Thijs L, Staessen JA, Clement DL, De Buyzere ML, De Bacquer DA. Night–day blood pressure ratio and dipping pattern as predictors of death and cardiovascular events in hypertension. Journal of Human Hypertension. 2009;23:645–653. doi: 10.1038/jhh.2009.9

39. McGonagle K, Freedman VA. The Panel Study of Income Dynamics’ Childhood Retrospective Circumstances Study (PSID-CRCS) User Guide: Final Release 1. In: Online: Institute for Social Research; 2015.

40. Kind AJH, Jencks S, Brock J, Yu M, Bartels C, Ehlenbach W, Greenberg C, Smith M. Neighborhood Socioeconomic Disadvantage and 30-Day Rehospitalization. Annals of Internal Medicine. 2014;161:765. doi: 10.7326/m13-2946

41. Knighton AJ, Savitz L, Belnap T, Stephenson B, Vanderslice J. Introduction of an Area Deprivation Index Measuring Patient Socio-economic Status in an Integrated Health System: Implications for Population Health. eGEMs (Generating Evidence & Methods to improve patient outcomes). 2016;4:9. doi: 10.13063/2327-9214.1238

42. Karaca A, Demirci N, Yılmaz V, Hazır Aytar S, Can S, Ünver E. Validation of the ActiGraph wGT3X-BT Accelerometer for Step Counts at Five Different Body Locations in Laboratory Settings. Measurement in Physical Education and Exercise Science. 2022;26:63–72. doi: 10.1080/1091367x.2021.1948414

43. Watso JC, Robinson AT, Babcock MC, Migdal KU, Witman MAH, Wenner MM, Stocker SD, Farquhar WB. Short-term water deprivation attenuates the exercise pressor reflex in older female adults. Physiological Reports. 2020;8. doi: 10.14814/phy2.14581

44. Harnack LJ, Cogswell ME, Shikany JM, Gardner CD, Gillespie C, Loria CM, Zhou X, Yuan K, Steffen LM. Sources of Sodium in US Adults From 3 Geographic Regions. Circulation. 2017;135:1775–1783. doi: 10.1161/circulationaha.116.024446

45. Lakens D. Calculating and reporting effect sizes to facilitate cumulative science: a practical primer for t-tests and ANOVAs. Front Psychol. 2013;4:863. doi: 10.3389/fpsyg.2013.00863

46. Kerby DS. The Simple Difference Formula: An Approach to Teaching Nonparametric Correlation. Comprehensive Psychology. 2014;3:11.IT.13.11. doi: 10.2466/11.it.3.1

47. Profant J, Dimsdale JE. Race and Diurnal Blood Pressure Patterns. Hypertension. 1999;33:1099–1104. doi: 10.1161/01.hyp.33.5.1099

48. Yano Y, Reis JP, Tedla YG, Goff DC, Jacobs DR, Sidney S, Ning H, Liu K, Greenland P, Lloyd-Jones DM. Racial Differences in Associations of Blood Pressure Components in Young Adulthood With Incident Cardiovascular Disease by Middle Age. JAMA Cardiology. 2017;2:381. doi: 10.1001/jamacardio.2016.5678

49. Chase HP, Garg SK, Icaza G, Carmain JA, Walravens CF, Marshall G. 24-h ambulatory blood pressure monitoring in healthy young adult Anglo, Hispanic, and African-American subjects. Am J Hypertens. 1997;10:18–23. doi: 10.1016/s0895-7061(96)00260-9

50. Harshfield GA, Pulliam DA, Somes GW, Alpert BS. Ambulatory blood pressure patterns in youth. Am J Hypertens. 1993;6:968–973. doi: 10.1093/ajh/6.11.968

51. Harshfield GA, Alpert BS, Willey ES, Somes GW, Murphy JK, Dupaul LM. Race and gender influence ambulatory blood pressure patterns of adolescents. Hypertension. 1989;14:598–603. doi: 10.1161/01.hyp.14.6.598

52. Muntner P, Lewis CE, Diaz KM, Carson AP, Kim Y, Calhoun D, Yano Y, Viera AJ, Shimbo D. Racial Differences in Abnormal Ambulatory Blood Pressure Monitoring Measures: Results From the Coronary Artery Risk Development in Young Adults (CARDIA) Study. American Journal of Hypertension. 2015;28:640–648. doi: 10.1093/ajh/hpu193

53. Wang X, Poole JC, Treiber FA, Harshfield GA, Hanevold CD, Snieder H. Ethnic and Gender Differences in Ambulatory Blood Pressure Trajectories. Circulation. 2006;114:2780–2787. doi: 10.1161/circulationaha.106.643940

54. Belsha C, Spencer H, Berry P, Plummer J, Wells T. Diurnal blood pressure patterns in normotensive and hypertensive children and adolescents. Journal of Human Hypertension. 1997;11:801–806. doi: 10.1038/sj.jhh.1000553

55. Falkner B, Lurbe E. Primordial Prevention of High Blood Pressure in Childhood. Hypertension. 2020;75:1142–1150. doi: 10.1161/hypertensionaha.119.14059

56. Firebaugh G, Acciai F. For blacks in America, the gap in neighborhood poverty has declined faster than segregation. Proceedings of the National Academy of Sciences. 2016;113:13372–13377. doi: 10.1073/pnas.1607220113

57. Mayne SL, Moore KA, Powell-Wiley TM, Evenson KR, Block R, Kershaw KN. Longitudinal Associations of Neighborhood Crime and Perceived Safety With Blood Pressure: The Multi-Ethnic Study of Atherosclerosis (MESA). Am J Hypertens. 2018;31:1024–1032. doi: 10.1093/ajh/hpy066

58. Mujahid MS, Gao X, Tabb LP, Morris C, Lewis TT. Historical redlining and cardiovascular health: The Multi-Ethnic Study of Atherosclerosis. Proceedings of the National Academy of Sciences. 2021;118:e2110986118. doi: 10.1073/pnas.2110986118

59. Kershaw KN, Robinson WR, Gordon-Larsen P, Hicken MT, Goff DC, Carnethon MR, Kiefe CI, Sidney S, Diez Roux AV. Association of Changes in Neighborhood-Level Racial Residential Segregation With Changes in Blood Pressure Among Black Adults. JAMA Internal Medicine. 2017;177:996. doi: 10.1001/jamainternmed.2017.1226

60. Snyder-Mackler N, Burger JR, Gaydosh L, Belsky DW, Noppert GA, Campos FA, Bartolomucci A, Yang YC, Aiello AE, O’Rand A, et al. Social determinants of health and survival in humans and other animals. Science. 2020;368. doi: 10.1126/science.aax9553

61. Merkin SS, Basurto-Dávila R, Karlamangla A, Bird CE, Lurie N, Escarce J, Seeman T. Neighborhoods and cumulative biological risk profiles by race/ethnicity in a national sample of U.S. adults: NHANES III. Ann Epidemiol. 2009;19:194–201. doi: 10.1016/j.annepidem.2008.12.006

62. Pretty C, O’Leary DD, Cairney J, Wade TJ. Adverse childhood experiences and the cardiovascular health of children: a cross-sectional study. BMC Pediatrics. 2013;13:208. doi: 10.1186/1471-2431-13-208

63. Saffer H, Dave D, Grossman M, Ann Leung L. Racial, Ethnic, and Gender Differences in Physical Activity. Journal of Human Capital. 2013;7:378–410. doi: 10.1086/671200

64. Kell KP, Judd SE, Pearson KE, Shikany JM, Fernández JR. Associations between socio-economic status and dietary patterns in US black and white adults. British Journal of Nutrition. 2015;113:1792–1799. doi: 10.1017/s0007114515000938

65. Garcia MT, Sato PM, Trude ACB, Eckmann T, Steeves ETA, Hurley KM, Bógus CM, Gittelsohn J. Factors Associated with Home Meal Preparation and Fast-Food Sources Use among Low-Income Urban African American Adults. Ecology of Food and Nutrition. 2018;57:13–31. doi: 10.1080/03670244.2017.1406853

66. Appel LJ, Moore TJ, Obarzanek E, Vollmer WM, Svetkey LP, Sacks FM, Bray GA, Vogt TM, Cutler JA, Windhauser MM, et al. A Clinical Trial of the Effects of Dietary Patterns on Blood Pressure. New England Journal of Medicine. 1997;336:1117–1124. doi: 10.1056/nejm199704173361601

67. Gohar EY, De Miguel C, Obi IE, Daugherty EM, Hyndman KA, Becker BK, Jin C, Sedaka R, Johnston JG, Liu P, et al. Acclimation to a High-Salt Diet Is Sex Dependent. Journal of the American Heart Association. 2022;11. doi: 10.1161/jaha.120.020450

68. Jackson RW, Treiber FA, Harshfield GA, Waller JL, Pollock JS, Pollock DM. Urinary excretion of vasoactive factors are correlated to sodium excretion. Am J Hypertens. 2001;14:1003–1006. doi: 10.1016/s0895-7061(01)02169-0

69. Hwang YS, Hsieh TJ, Lee YJ, Tsai JH. Circadian rhythm of urinary endothelin-1 excretion in mild hypertensive patients. Am J Hypertens. 1998;11:1344–1351. doi: 10.1016/s0895-7061(98)00170-8

70. Chrysant SG, Danisa K, Kem DC, Dillard BL, Smith WJ, Frohlich ED. Racial differences in pressure, volume and renin interrelationships in essential hypertension. Hypertension. 1979;1:136–141. doi: 10.1161/01.hyp.1.2.136

71. Rifkin DE, Khaki AR, Jenny NS, McClelland RL, Budoff M, Watson K, Ix JH, Allison MA. Association of renin and aldosterone with ethnicity and blood pressure: the Multi-Ethnic Study of Atherosclerosis. Am J Hypertens. 2014;27:801–810. doi: 10.1093/ajh/hpt276

72. Stanhewicz AE, Wong BJ. Counterpoint: Investigators should not control for menstrual cycle phase when performing studies of vascular control that include women. J Appl Physiol (1985). 2020;129:1117–1119. doi: 10.1152/japplphysiol.00427.2020

73. Williamson PM, Buddle ML, Brown MA, Whitworth JA. Ambulatory blood pressure monitoring (ABPM) in the normal menstrual cycle and in women using oral contraceptives. Comparison with conventional blood pressure measurement. Am J Hypertens. 1996;9:953–958. doi: 10.1016/0895-7061(96)00150-1

## Supplemental Reference

1. Nichols OI, Fuller-Rowell TE, Robinson AT, Eugene D, Homandberg LK. Neighborhood Socioeconomic Deprivation in Early Childhood Mediates Racial Disparities in Blood Pressure in a College Student Sample. Journal of Youth and Adolescence. 2022;51:2146–2160. doi: 10.1007/s10964-022-01658-6

2. Williams DR, Yu Y, Jackson JS, Anderson NB. Racial Differences in Physical and Mental Health. Journal of Health Psychology. 1997;2:335–351. doi: 10.1177/135910539700200305

3. Krieger N, Smith K, Naishadham D, Hartman C, Barbeau EM. Experiences of discrimination: Validity and reliability of a self-report measure for population health research on racism and health. Social Science & Medicine. 2005;61:1576–1596. doi: 10.1016/j.socscimed.2005.03.006

4. Felitti VJ, Anda RF, Nordenberg D, Williamson DF, Spitz AM, Edwards V, Koss MP, Marks JS. Relationship of childhood abuse and household dysfunction to many of the leading causes of death in adults. The Adverse Childhood Experiences (ACE) Study. Am J Prev Med. 1998;14:245–258. doi: 10.1016/s0749-3797(98)00017-8

5. Cronholm PF, Forke CM, Wade R, Bair-Merritt MH, Davis M, Harkins-Schwarz M, Pachter LM, Fein JA. Adverse Childhood Experiences: Expanding the Concept of Adversity. Am J Prev Med. 2015;49:354–361. doi: 10.1016/j.amepre.2015.02.001

6. Finkelhor D, Shattuck A, Turner H, Hamby S. Improving the adverse childhood experiences study scale. JAMA Pediatr. 2013;167:70–75. doi: 10.1001/jamapediatrics.2013.420

7. Knaier R, Höchsmann C, Infanger D, Hinrichs T, Schmidt-Trucksäss A. Validation of automatic wear-time detection algorithms in a free-living setting of wrist-worn and hip-worn ActiGraph GT3X. BMC Public Health. 2019;19:244. doi: 10.1186/s12889-019-6568-9

8. Montoye AHK, Clevenger KA, Pfeiffer KA, Nelson MB, Bock JM, Imboden MT, Kaminsky LA. Development of cut-points for determining activity intensity from a wrist-worn ActiGraph accelerometer in free-living adults. J Sports Sci. 2020;38:2569–2578. doi: 10.1080/02640414.2020.1794244

9. Marino M, Li Y, Rueschman MN, Winkelman JW, Ellenbogen JM, Solet JM, Dulin H, Berkman LF, Buxton OM. Measuring Sleep: Accuracy, Sensitivity, and Specificity of Wrist Actigraphy Compared to Polysomnography. Sleep. 2013;36:1747–1755. doi: 10.5665/sleep.3142

10. Chow CM, Wong SN, Shin M, Maddox RG, Feilds KL, Paxton K, Hawke C, Hazell P, Steinbeck K. Defining the rest interval associated with the main sleep period in actigraph scoring. Nat Sci Sleep. 2016;8:321–328. doi: 10.2147/nss.s114969

11. Fuller-Rowell TE, Nichols OI, Robinson AT, Boylan JM, Chae DH, El-Sheikh M. Racial disparities in sleep health between Black and White young adults: The role of neighborhood safety in childhood. Sleep Med. 2021;81:341–349. doi: 10.1016/j.sleep.2021.03.007

12. Culver MN, Mcmillan NK, Cross BL, Robinson AT, Montoye AH, Riemann BL, Flatt AA, Grosicki GJ. Sleep duration irregularity is associated with elevated blood pressure in young adults. Chronobiology International. 2022:1–9. doi: 10.1080/07420528.2022.2101373

13. Hoopes EK, Berube FR, D’Agata MN, Patterson F, Farquhar WB, Edwards DG, Witman MAH. Sleep duration regularity, but not sleep duration, is associated with microvascular function in college students. Sleep. 2021;44. doi: 10.1093/sleep/zsaa175

14. Food Amount Reporting Booklets. Nutrition Coordinating Center (NCC), University of Minnesota. Accessed October 4th.

